# Investigating heterogeneous genetic effects contributing to smoking behaviors

**DOI:** 10.1101/2021.08.08.21261764

**Authors:** Scott A Funkhouser, Jason D Boardman, John K Hewitt, Michael C Stallings, Christian J Hopfer, Sandra A Brown, Tamara L Wall, Chandra A Reynolds, Matthew C Keller, Luke M Evans

## Abstract

The genetic architecture of numerous smoking behaviors is highly polygenic, but these genetic effects are heterogeneous and depend on moderating factors. Here, we used common SNPs from the UK Biobank to investigate heterogeneous genetic effects for smoking heaviness using cigarettes per day (CPD) records, smoking initiation (SI), and smoking cessation (SC). We assessed heterogeneous effects across levels of sex, age of smoking initiation, major depressive disorder (MDD) DSM-V-like diagnosis, generalized anxiety disorder (GAD) DSM-V-like diagnosis, and whether an individual has seen a psychiatrist for nerves, anxiety, tension, or depression. We observed suggestive evidence of heterogeneous genetic effects for CPD and SC, moderated by MDD and GAD, respectively [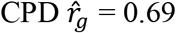 (SE = 0.15) between MDD cases and controls, and 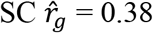 (SE = 0.28) between GAD cases and controls, *p* < 0.05]. We detected 5 SNPs with genome-wide significant evidence of heterogeneous effects moderated by either MDD or GAD (*p*-value < 5×10^-8^) for CPD and SC. We also observed strong evidence for heterogeneous genetic effects for SI between sexes (between-sex 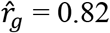, SE = 0.02). While we detected no individual SNPs that were moderated by sex at genome-wide significance (all *p*-value > 5×10^-8^), we observed evidence of novel genome-wide significant SI-SNP associations using sex-stratified GWAS; six loci were discovered in either men or women separately that were not identified in a previous smoking meta-analysis that had a 6-fold larger, sex-combined sample. Furthermore, using several independent testing samples, there was suggestive evidence that the prediction ability of a polygenic risk score (PRS) for smoking initiation improved through the utilization of sex-specific SNP effects. This work suggests that a more nuanced approach to GWAS analyses is warranted, as potential heterogeneous effects can complicate variant discovery and polygenic risk score accuracy.

**Author Summary:** Smoking imposes a heavy health burden and is highly polygenic in architecture. Consistent with most complex traits, many causal loci have yet to be identified, even when using the largest available samples. One possible reason for the difficulty in inferring genetic variants associated with such complex traits is that common genetic variants possess context-dependent (or heterogeneous) effects. Utilizing the UK Biobank, we find evidence for heterogeneous SNP effects on smoking initiation, heaviness, and cessation among psychiatric disorder cases and controls and between sexes. Failure to model such heterogeneity (when accounting for sample size) resulted in lower independent sample predictive ability. This work encourages a more nuanced approach to GWAS and polygenic risk prediction. The assumption that all genetic effects are homogeneous limits our understanding of complex traits when heterogeneous effects are present.

## Introduction

Tobacco smoking has contributed to more than 20 million preventable deaths since 1964[1] and disproportionally affects certain groups within populations. Individuals with depression and anxiety are at an increased risk of becoming nicotine dependent[2–4] and early-onset smokers are more likely to smoke heavily compared to late-onset smokers[5]. Furthermore, sex differences in smoking behaviors are well-documented, with a consensus showing men exhibit greater rates of smoking initiation, cessation, and tobacco usage than women[6–8]. While numerous environmental factors likely contribute to tobacco’s heterogeneous usage, smoking behaviors are known to be heritable[9,10] with a highly polygenic architecture comprised of many loci with very small effects[11–15]. Family studies have suggested that the genetic component to smoking is itself heterogeneous[16–19], *i*.*e*. dependent on certain factors that moderate one’s genetic risk to smoke, smoke heavily, or persist in smoking.

Heterogeneous genetic effects could complicate efforts to identify genetic loci that influence smoking behaviors. While many genome-wide significant (GWS; *p*-value < 5×10^-8^) SNPs have been identified, their individual effects are small and collectively explain only about one-third of the SNP-heritability[20]. If many of the genetic effects for smoking are heterogeneous, this could partially explain i) differing tobacco usage across groups and ii) the exceedingly small size of average SNP effects when modeled as having a single effect across all groups or conditions. This second point may be especially true when the effect of an allele takes place only in a rare group (*e*.*g*., psychiatric disorder cases), where the average effect of an allele substitution (*i*.*e*., averaged across a random sample of psychiatric disorder cases and controls) would be weighted toward zero. Prior evidence from a Japanese population suggests sex-dependent SNP effects for smoking behaviors[13], but few studies have utilized genome-wide SNP data to infer heterogeneous genetic effects for smoking behaviors across potential moderating factors. Other candidate gene studies have implicated individual loci with heterogeneous smoking effects[21], but these findings have not replicated in biobank-scale data[22].

In this study, we estimated heterogeneous genetic effects for smoking heaviness [*i*.*e*., cigarettes per day (CPD)], smoking cessation (SC), and smoking initiation (SI) using the UK Biobank[23]. Heterogeneity of effects was assessed across levels of sex, age of smoking initiation (ASI), major depressive disorder (MDD) DSM-V-like diagnosis, generalized anxiety disorder (GAD) DSM-V-like diagnosis, and whether an individual had seen a psychiatrist for nerves, anxiety, tension, or depression (PSYCH). We evaluated evidence for heterogeneous genetic effects at multiple scales: genome-wide, within functional annotations, and at single SNPs. Specifically, for each smoking trait we i) estimated the genetic correlation between groups (*e*.*g*., between MDD cases and controls) and the proportion of phenotypic variance explained by heterogeneous SNP effects genome-wide, ii) estimated differences in heritability enrichment among cell-type-specific annotations, and iii) conducted a GWAS to test for heterogeneous effects at individual SNPs. Given clear genome-wide evidence for differential genetic architectures for SI between sexes (as evident from a between-sex 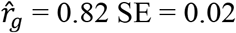), we then evaluated polygenic risk score (PRS) accuracy for SI when allowing for SNP effects to be heterogeneous, as opposed to homogenous, between sexes.

## Results

### Clear genome-wide evidence of heterogenous SNP effects for SI between sexes

Using common (MAF > 0.01) genome-wide SNPs, we modeled each smoking trait (CPD, SI, and SC) across levels of potential moderators (*e*.*g*., across males and females or MDD-like cases and controls) using a bivariate GREML model (see Methods). CPD is often transformed to different scales prior to analysis[16,21]; because SNP effect heterogeneity may depend on the chosen scale, we considered four different transformations of CPD for all analyses: the raw scale, binned, log transformed, and dichotomized (see Methods for more details). For all trait-by-moderator combinations, we restricted our analyses to unrelated individuals (estimated relatedness <0.05, sample sizes in Table 1).

**Table 1.**
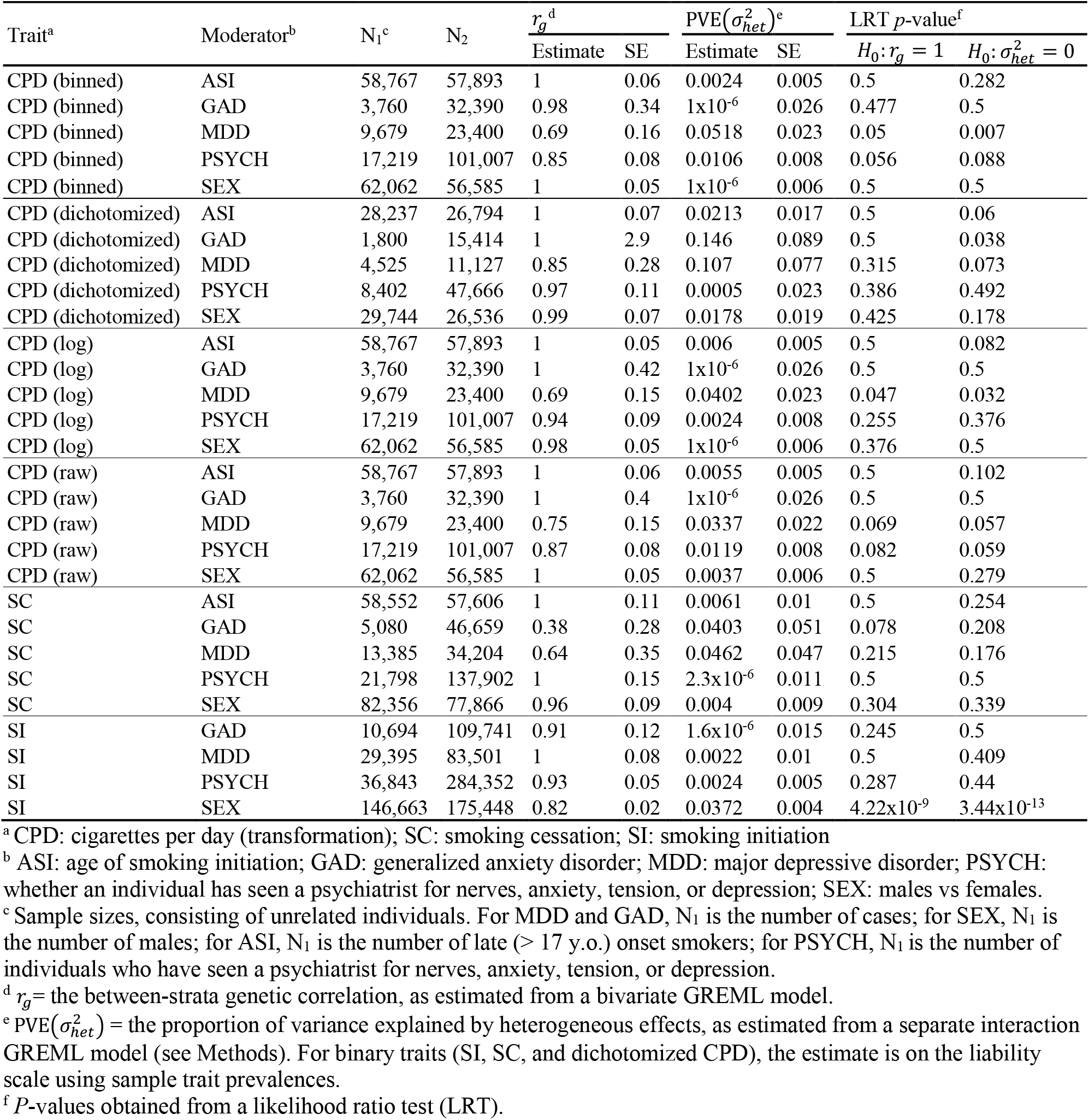
Genome-wide estimates of heterogeneous genetic effects

From the bivariate model we estimated the genetic correlation (*r*_*g*_) between strata to infer the presence of heterogeneous genetic effects that show disproportionality between strata (Table 1). Using a likelihood ratio test under the null hypothesis of *r*_*g*_ = 1, we found limited evidence of different genetic effects across most moderators. However, we found strong evidence that genetic effects for SI differed between sexes (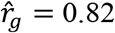, SE = 0.02; *p*-value = 4.22 × 10^-9^), and nominally significant evidence that genetic effects for log-transformed CPD differed between MDD-like cases and controls (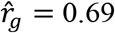, SE = 0.15; *p*-value = 0.047). We also found suggestive evidence that genetic effects for SC differed between GAD-like cases and controls, but due to the relatively small number of GAD cases, the standard error was quite large (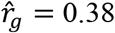, SE = 0.28; *p*-value = 0.078). Estimates of genetic correlations using an alternative method, cross-trait LDSC[24], were consistent with GREML-based estimates (Supplementary Table S1).

We next decomposed the total variance across strata to estimate the proportion of phenotypic variance explained by heterogeneous effects [denoted 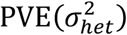] using a GREML interaction model[25] (Table 1). For SI, we estimated sex-dependent effects to account for 3.7% of the liability scale phenotypic variance (SE = 0.4%; *p*-value = 3.44×10^-13^). Likewise, for binned CPD, we estimated MDD-dependent genetic effects to account for 5.2% of the total variance but with larger uncertainty (SE = 2.3%; *p-*value = 0.007).

To determine if genetic variances themselves differed between strata, we estimated fold differences in variance components as estimated from the bivariate model (Fig. 1). For CPD (binned, dichotomized, and raw scales), we observed nominal evidence of differing genetic variances between males and females, and between late-onset smokers and early-onset smokers (95% confidence interval of 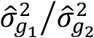 did not include 1). No other smoking phenotype showed evidence of differing genetic variances between moderator groups. Given that allele frequencies between strata are highly correlated (the genome-wide correlations of allele frequencies between strata were all >0.999), we hypothesized that differing genetic variances may reflect SNP effects that depend on the trait variance itself, for instance, the greater total variance of CPD in males than females (Fig 1). We repeated the bivariate analysis after standardizing the trait within strata (centering to zero mean and scaling to unit variance) and observed no clear differences in genetic or residual variances between strata. This indicates that while SNP effects for CPD may differ between sexes or between late-onset and early-onset smokers, such differences in SNP effects for CPD can be accounted for by differences in trait variance rather than differing genetic mechanisms.

**Fig 1.**
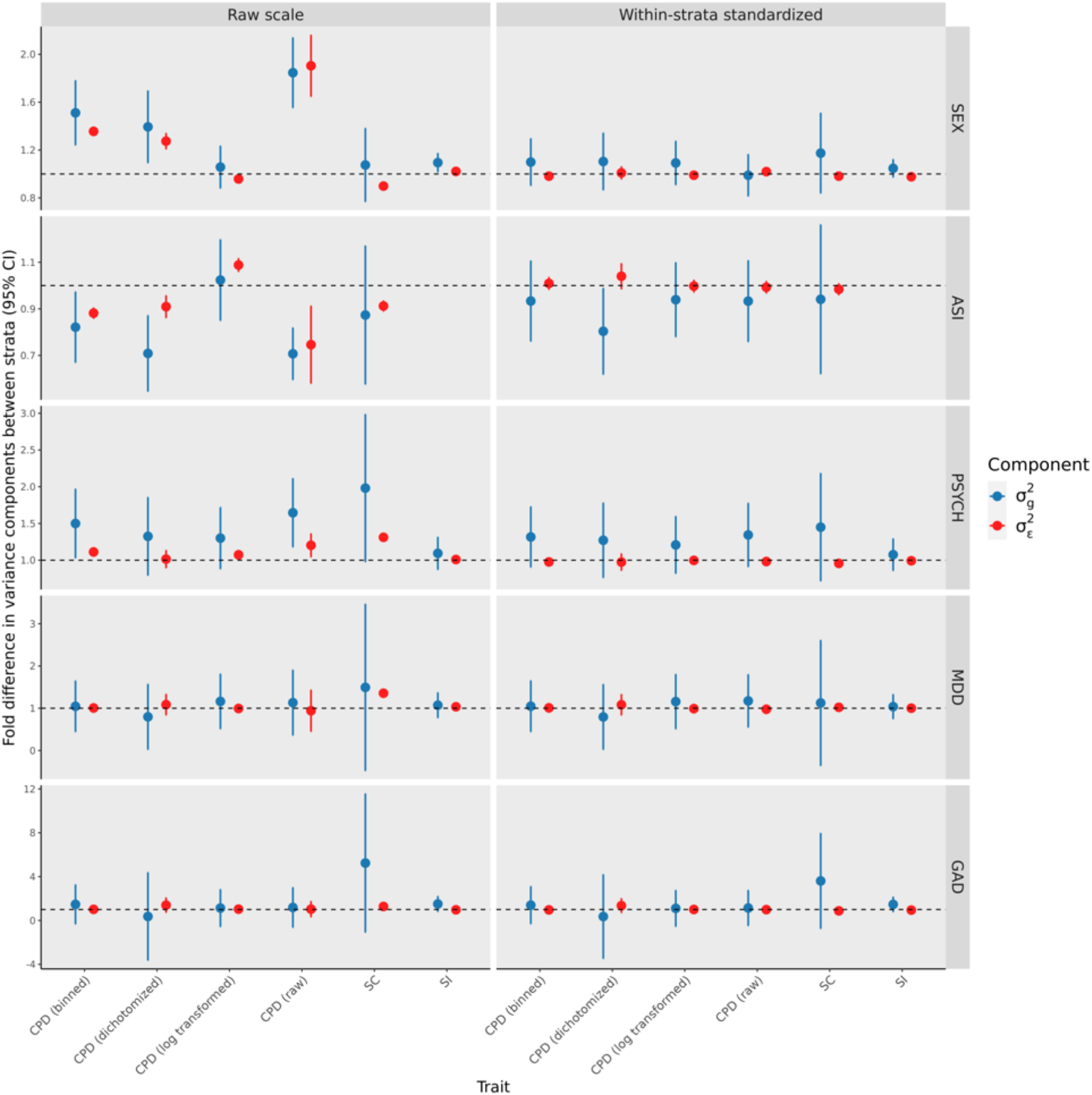
Fold differences in variance components between strata. Shown are point estimates and 95% confidence intervals. The dashed horizontal line at 1 indicates equal variance components. Each horizontal facet indicates a different moderator. For ASI, larger values than 1 indicate larger variances in late onset (Age 17-35) smokers than early onset smokers (Age 10-16). For both GAD and MDD, larger values than 1 indicate larger variances in cases than controls. For PSYCH, larger values than 1 indicate larger variances in those who have seen a psychiatrist for nerves than those who have not. For SEX, larger values than 1 indicate larger variances among males than females. For “Raw scale” the bivariate model is fit without standardizing the trait. For “Within-strata standardized”, the bivariate model is fit after centering and scaling the trait within strata.

### No evidence of differing partitioned heritabilities within cell-type-specific annotations

Given evidence for disproportional genome-wide genetic effects, particularly for SI between sexes, we next asked whether evidence for differing SNP effects can be localized to functional genomic annotations. We performed stratified (*e*.*g*., by sex) GWAS using BOLT-LMM[26], then used LD score regression (LDSC)[27,28] to partition the SNP-based heritability (*h*^*2*^_*SNP*_) within each strata, estimating strata-specific LDSC regression coefficients and *h*^*2*^_*SNP*_ enrichment scores. When dichotomizing CPD, we were unable to use BOLT-LMM with GAD cases due to a limited sample size (N = 1854; full BOLT-LMM sample sizes shown in Supplementary Table S2) and therefore were unable to compare partitioned heritabilities for dichotomized CPD between GAD cases and controls. We tested a total of 221 annotations derived from four gene expression datasets[28], with each annotation corresponding to a set of SNPs within 100-kb of genes uniquely expressed in a particular tissue. We used a z-score to test for differences in LDSC coefficients and *h*^*2*^_*SNP*_ enrichment scores between strata (see Methods). Across all 6188 tests (221 annotations by 28 trait-by-moderator combinations), we found no significant differences in LDSC regression coefficients or *h*^*2*^_*SNP*_ enrichment scores after controlling for the false-discovery rate (Supplementary Figures S1-S2), indicating no evidence that groups differ in the heritable contribution of cell-type specific or other functional annotations.

### Individual SNPs show evidence of heterogeneous effects

We next tested for differences in marginal SNP effect estimates between strata using a z-score of the difference in effect sizes and a two-sided *p*-value (*p*-diff; see methods; Supplementary Figures S3-S58). Across all trait-by-moderator combinations tested, we observed 5 loci with genome-wide significant (GWS) evidence of heterogenous effects (*p-*diff < 5×10^-8^; Table 2). Notably, all 5 loci showed differing directions of effects of lead SNPs between strata, and none of these loci reached genome-wide significance within strata. Miami-plots that compare within-strata GWAS results can be found in Supplementary Figures S59-S86.

**Table 2.**
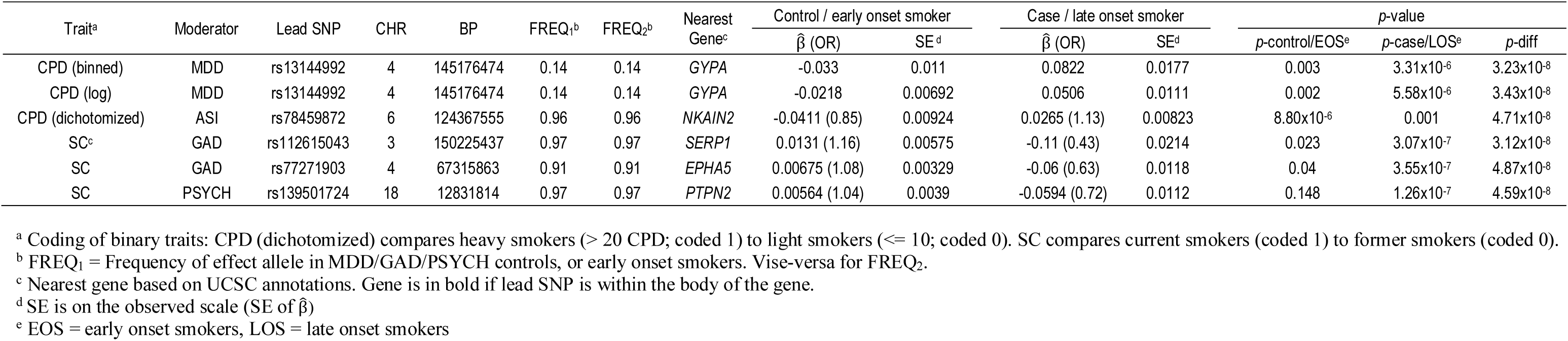
Genome-wide significant differences in SNP effects (*p-*diff < 5×10^-8^)

For CPD, SNPs with significant differences in effects between strata were largely dependent on the CPD transformation; both binned CPD and log transformed CPD showed GWS evidence of differing SNP effects at rs13144992 (*p*-diff = 3.23×10^-8^ and 3.43×10^-8^, respectively) between MDD cases and controls, however this signal decayed with raw CPD (*p*-diff = 1.4×10^-7^) and dichotomized CPD (*p*-diff = 1.89×10^-4^). Likewise, when dichotomizing CPD, the only heterogenous signal reaching GWS was between late-onset and early-onset smokers, located at rs78459872, a SNP that exhibited no GWS evidence of ASI-dependent effects under different CPD transformations (*p*-diff > 4.3×10^-5^). We further observed two loci with differing effects for SC between GAD cases and controls, and one locus with differing SC effects between PSYCH cases and controls. Given evidence that several traits possess differences in variance between strata (Fig. 1), we re-tested these five SNPs after standardizing the corresponding trait within strata to see if SNP effect differences may reflect differences in trait scale (Supplementary Table S3). After normalizing and re-testing, we found very little differences in *p*-diff values, however both the ASI-dependent SNP associated with dichotomized CPD (rs78459872) and the PSYCH- dependent SNP associated with SC (rs139501724) were no longer GWS (both *p*-diff = 5.3×10^-8^ after re-testing). Additional loci reached GWS in one stratum but not the other, however most of these instances (*e*.*g*. a GWS signal in MDD-controls but not in MDD-cases) are likely explained by differences in power (see Supplementary Figures S59-S86).

### Novel SI-associated loci detected using sex-stratified GWAS

We found strong genome-wide evidence of heterogeneous genetic effects for smoking initiation between sexes, consistent with evidence from an independent Japanese sample [13]. However, we observed no sex differences in individual SNP effects for SI at GWS (see Supplementary Figures S57-S58). Due to this surprising lack of genome-wide significant effect differences at individual SNPs, we performed additional analyses to characterize the genetic architecture of SI between sexes. Genome-wide, we observed an inverse relationship between MAF and estimated sex-differences in SNP effects, and simulations indicated a lack of power to detect GWS SNP effect differences across the MAF spectrum (Supplementary Figure S87).

Despite the lack of power to detect significant differences in effect estimates, our sample was well-powered to detect effects in either sex alone. In the sex-stratified GWAS for SI, we observed 51 GWS risk loci, consisting of 24 male risk loci not overlapping with a female risk locus, 23 female risk loci not overlapping with a male risk locus, and 4 risk loci that overlapped between males and females (Fig 2; Supplementary Tables S4 and S5). To determine whether novel SI-associated loci may be identified through sex-stratified GWAS, in which genetic effects are allowed to be heterogeneous between sexes, we compared our sex-stratified GWAS results to sex-combined GWAS results, in which a single effect is assumed to be shared by sexes (N = 418,329). We observed 14 independent (r^2^ < 0.1) lead SNPs using sex-stratified GWAS that were not within a risk locus identified from the sex-combined GWAS, despite the roughly 2-fold larger sample size of the sex-combined analysis. Furthermore, six of these 14 loci were also not detected in a prior European ancestry meta-analysis of SI (the trait definition was identical to this study, see Methods for more details) involving roughly 6-fold more individuals than either sex-stratified analysis (N ∼ 1.2 million[11], Fig 2 and Table 3). To quantify the degree that sex-stratified GWAS can lead to increased statistical power in detecting loci bearing heterogenous effects, we performed power analyses, varying sample size, MAF, and degree of SNP effect heterogeneity (Supplementary Figure S88). Sex-stratified GWAS consistently showed equal or greater power to detect any effect (whether it affects males, females, or both) at a sex-heterogeneous effect locus than sex-combined GWAS. For example, we observed 3-fold increase in power at somewhat rare SNPs (MAF = 0.05) where the fold-difference in sex-specific odds ratios was 1.04; when observing real data, we found such differences in effects at MAF = 0.05 to be within a plausible range, indicating that our present sample sizes are simply underpowered to detect true heterogeneous effects between sexes (see Supplementary Figure S87).

**Fig 2.**
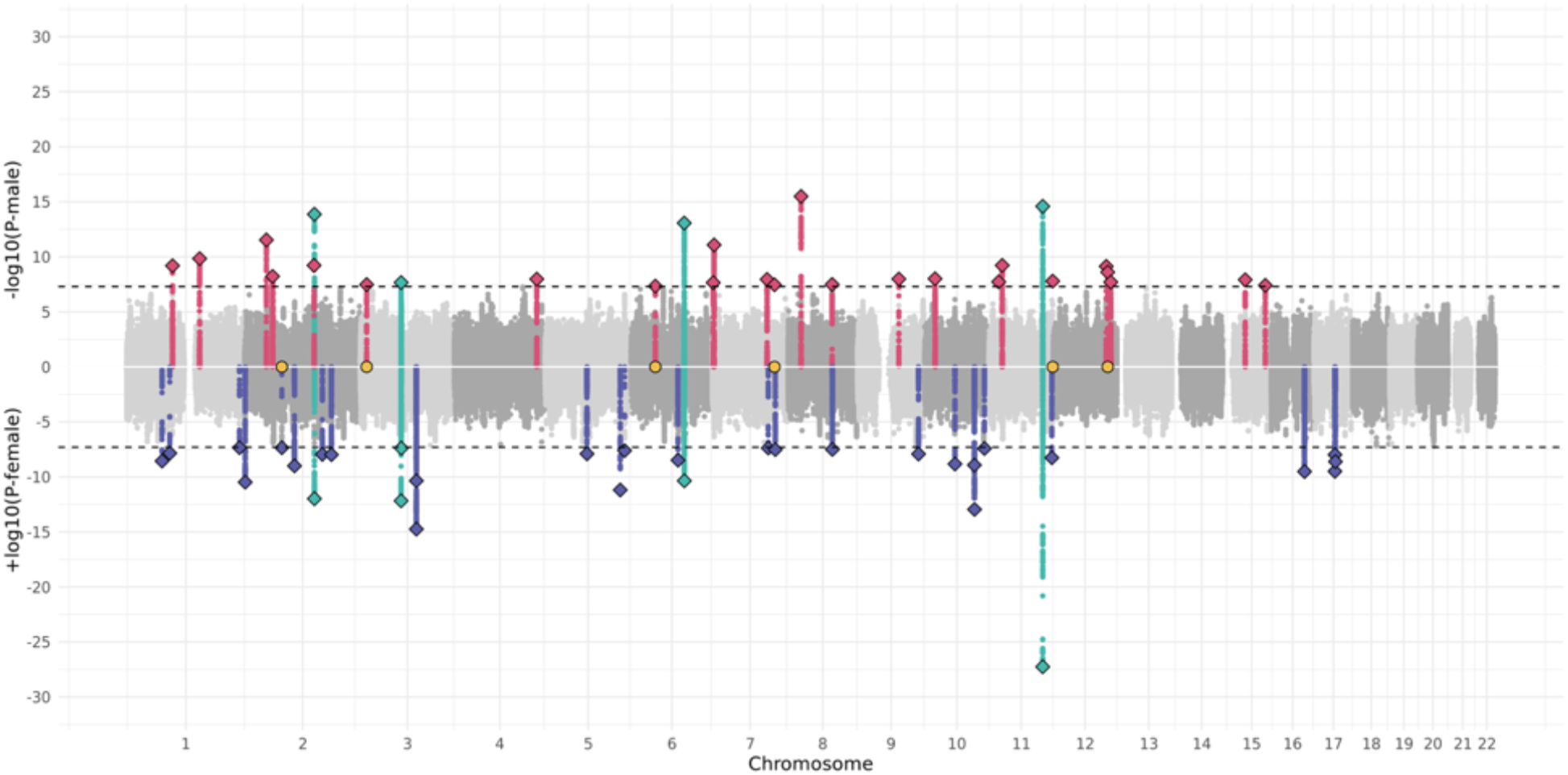
Miami-plot showing sex-specific GWAS signals for smoking initiation. The male manhattan plot for SI is shown above 0 on the x-axis, while the female manhattan plot for SI is shown below 0 on the x-axis. SNPs are highlighted red if they were within a male-specific genomic risk locus (a locus that did not overlap with any female-specific GWS risk locus), and vice versa for blue SNPs. In teal are SNPs within risk loci that overlapped between males and females. Diamonds indicate independent (r^2^ < 0.1), sex-specific lead SNPs. Yellow circles mark the position of novel signals (Table 3)—lead SNPs that reached GWS in either males or females but were not within a detectable risk locus when performing a sex-combined GWAS in the UK Biobank nor within a risk locus in a prior meta-analysis of SI (N ∼ 1.2M)[11].

**Table 3.**
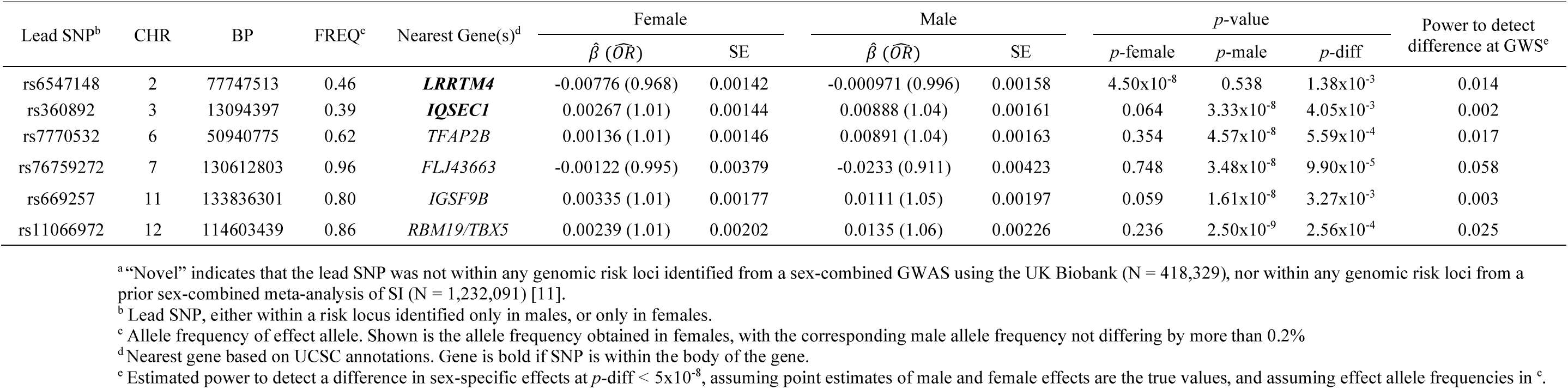
Novel^a^ SI-associated loci discovered using sex-stratified GWAS

For males and females separately, we then estimated the genetic correlation between SI and 757 sex-combined traits on LDhub [29]. Despite observing very different GWS signals between males and females for SI, we observed that for all 757 traits, the male-specific SI genetic correlation estimate (95% CI) overlapped with the female-specific SI genetic correlation estimate (95% CI) (Supplementary Tables S6-S7).

### Suggestive evidence of enhancing SI PRS accuracy using sex-specific SNP effects

Using independent target data from the National Longitudinal Study of Adolescent to Adult Health (Add Health) [30] and from the Center on Antisocial Drug Dependence (CADD) [31], we tested whether allowing for sex-specific SNP effects can enhance polygenic risk score (PRS) accuracy for SI, when compared to assuming SNP effects are shared between sexes. We computed male-specific and female-specific PRSs from the corresponding sex-stratified GWAS summary stats (derived from the UK Biobank) using SBLUP in GCTA [25]. For comparison, we computed PRS from the sex-combined GWAS summary statistics and a sex-combined GWAS in which sample size was halved to resemble the sex-stratified sample sizes. Additionally, we computed another PRS derived from weighted sex-specific GWAS statistics, as implemented in SMTpred[32]. We computed prediction accuracy of PRS when compared to a covariate-only model using logistic regression and Nagelkerke’s R^2^ (see Methods).

Prediction accuracy was similar between the two independent target datasets across all training methods. Prediction accuracy was highest using the full, sex-combined training sample (Fig. 3; Supplementary Table S8). Sex-stratification of the GWAS slightly improved prediction compared to the sex-combined analysis when the training sample sizes were approximately equal, but the prediction standard errors were large and the improvement was not statistically significant. Using a weighted index to borrow information across sex-specific SNP effects improved prediction accuracy to a level comparable to that obtained with the full, sex-combined training sample.

**Fig 3.**
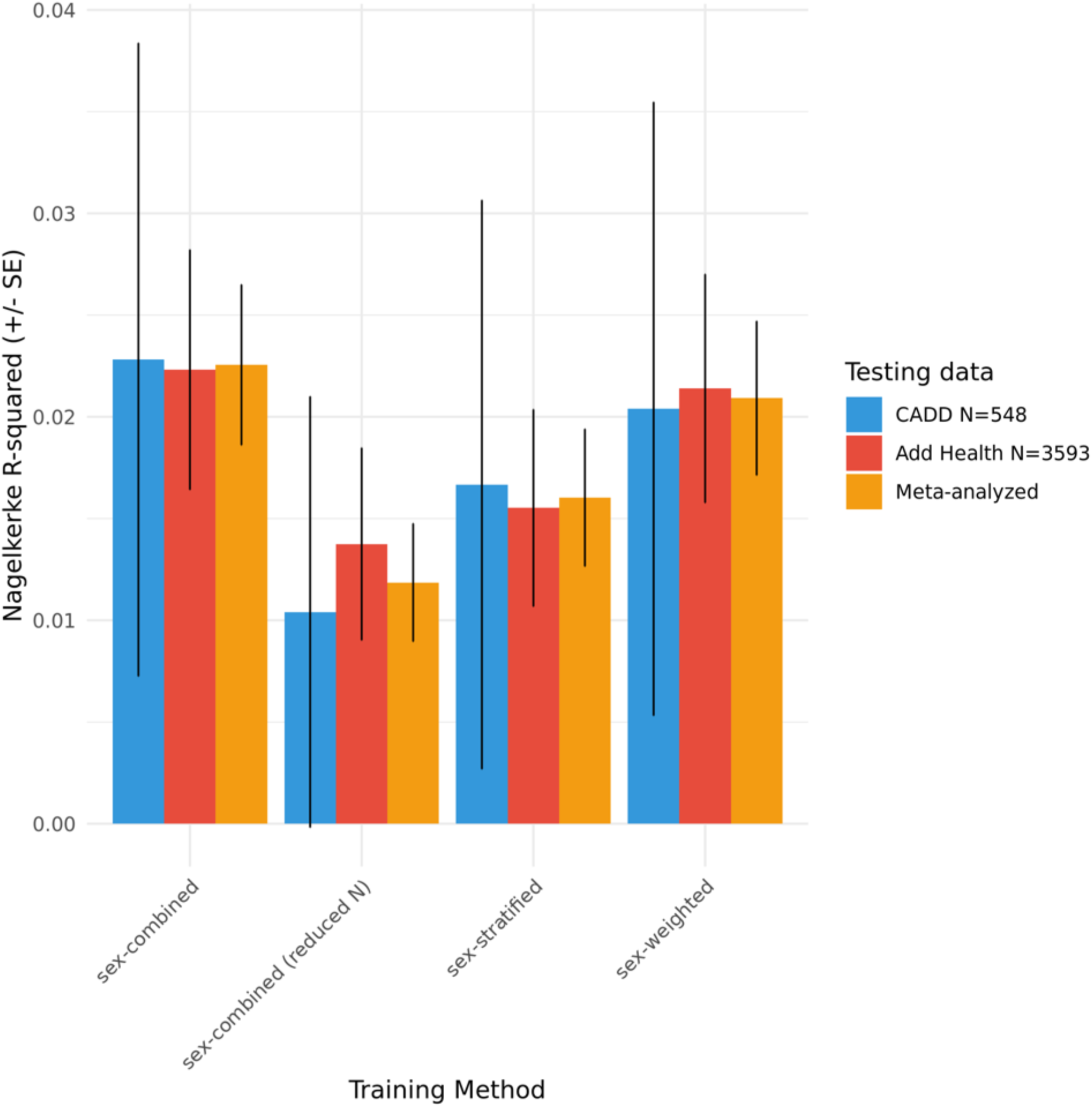
Accuracy of polygenic risk scores for smoking initiation using two testing datasets. Shown is the Nagelkerke R^2^ and intervals show one standard error obtained from 1000 bootstrap samples. Meta-analysis across datasets was done using inverse-variance weighting. Sex-combined (reduced N) used a reduced training sample size to roughly match the sample size obtained through sex-stratification. Sex-combined training N = 418,329; sex-combined (reduced N) training N = 209,157; sex-weighted and sex-stratified training Ns = 189693 males and 228636 females.

## Discussion

We observed modest evidence of genome-wide genetic moderation of smoking behaviors due to psychiatric disorders and age of smoking initiation, and several individual loci with differing strata-specific SNP effects. However, we observed strong genome-wide evidence that sex moderates the genetic contribution to smoking initiation, and identified numerous novel loci associated with SI when we stratified the GWAS by sex. These novel loci were not detected in a prior sex-combined GWAS of the same phenotype [11], despite utilizing a 6-fold increase in sample size than the sex-stratified GWAS presented in this study. This suggests that more nuanced analyses can aid in uncovering the genetic factors that contribute to the initiation of regular smoking and possibly in complex traits more generally.

### Psychiatric disorder moderation

Twin-based studies have found depression moderates the genetic variance of smoking heaviness[16]. We observed suggestive evidence that the genetic correlations of CPD between MDD cases and controls and SC between GAD cases and controls are less than one. A lack of stronger evidence is likely due to the low power to detect genetic variance differences between cases and controls (*e*.*g*., the number of unrelated GAD cases with an SC record was ∼5K, Table 1).

Consistent with these findings, we observed relatively few genome-wide significant SNP effects that differed among groups, with at least one depending on the trait scale. Within 100kb of these SNP-by-moderator associations are loci previously associated with COPD and lung cancer[33], forced vital capacity[34], waist circumference [35], forced vital capacity in COPD patients[36], sleep quality[37], and dinner intake in a Hispanic population[38]. Collectively, these results are consistent with a modest genetic effect moderation on smoking by depression and generalized anxiety, which is consistent with reports using twin samples, though of smaller magnitude[16–19]. Larger, independent samples with well-phenotyped psychiatric data—which are currently limited—will be necessary to identify heterogeneous effect loci, but we expect the difference in magnitude of individual effects will be small.

### Moderation by sex and age of smoking initiation

As seen in Fig. 1, depending on how one measures smoking heaviness (through different transformations of CPD), we observed different variances but no evidence of differential SNP heritabilities of CPD between males and females, and between late and early onset smokers. Non-genetic factors may largely be the direct cause of differing variances in CPD, with genetic effects differing proportionally due to these differences in scale. We did observe a single SNP with differing CPD effects between late-onset and early-onset smokers, however again this effect depended on the chosen CPD scale and transformation.

Alternatively, we observed clear evidence of disproportional genetic effects for SI between males and females, indicative of sex differences in genetic mechanisms that contribute to smoking initiation risk. This is consistent with a previous, independent study that estimated the between-sex genetic correlation to be < 1 for smoking initiation (measured as ever vs. never smokers) using common SNPs in a Japanese population[13]. Intriguingly, we identified no individual SNP effects at that differed between sexes at genome-wide significance (*p*-diff < 5×10^-8^), indicating that such effects are exceedingly small. Furthermore, we observed no evidence of differing genetic correlations with SI between males and females when testing over 700 traits on LD Hub. This could indicate that while males and females possess partially distinct genetic loci for SI, the functional consequences of each may not differ dramatically, genome-wide.

### Novel SI-associated loci identified through sex-stratified GWAS

While increasing sample size is one strategy to uncover novel risk loci for SI, the presence of heterogenous SNP effects could enable a more nuanced, sex-stratified analysis to uncover certain SI-associated loci more efficiently (see Fig 2; Supplementary Fig S88). We identified 6 GWS loci using sex-stratified GWAS for SI that were not detected in a sex-combined GWAS of the UK Biobank, nor detected in the largest known GWAS meta-analysis of SI[11]. These loci highlight the fact that, in addition to the improved power gained from increasing sample size such as in Liu et al.[11], incorporating nuanced analyses to investigate possible heterogeneous effects among groups can identify novel associations and provide a better understanding of the underlying trait architecture. Please see the Supplementary Note for discussion about these six novel signals.

A more nuanced association analysis may also improve genomic prediction. Leveraging sex-specific SNP effects for sex-stratified SI prediction appeared to increase accuracy when compared to sex-combined prediction using a comparable training sample size. When borrowing information between sex-specific SNP BLUPs using a weighted index as implemented in SMTpred[32], prediction accuracy increased to a similar level obtained when training with the full, sex-combined training sample. Future work may seek to develop additional means to borrow information between males and females to optimize prediction accuracy while allowing for heterogeneous genetic effects. Although the large standard errors complicate distinguishing the optimal approach among sex-combined, sex-weighted, and sex-stratified training methods, our results are consistent with improved prediction when allowing for heterogeneous effects, when accounting for training sample size.

## Limitations

Requiring individuals to possess both smoking and moderator phenotypes reduced our sample sizes, in some cases, severely (*e*.*g*., the aforementioned 5K individuals possessing both a SC and GAD record), which led to reduced power and greater uncertainty in effect estimates. For example, we detected GWS evidence of heterogeneous SNP effects associated with CPD and SC, however none of these SNPs reached GWS within strata. Furthermore, we cautiously note that sex-specific risk loci identified from two independent GWAS do not necessarily imply heterogeneous effects between sexes. In particular, if two non-overlapping risk loci are in close proximity, identification in one but not the other sex-specific GWAS may result from random sampling of genotypes rather than heterogeneous genetic effects, or from subtle differences in power (N_males_ = 189,693 and N_females_ = 228,636). Differential calling of genotypes or genotype sampling could partially explain why some sex-specific SNP effects reaching GWS in this study did not reach GWS in a prior meta-analyzed sample of the same phenotype. For studying the effects of MDD or GAD moderation, we emphasize the pressing need for an independent, replication dataset, however, currently there are very few samples containing large numbers of genotyped individuals with both smoking records and MDD-DSMV-like/GAD-DSMV-like records. Further work will be crucial in investigating MDD- and GAD-dependent genetic effects that contribute to smoking behaviors.

## Summary

For highly complex traits such as smoking behaviors, incorporating more nuanced analyses, such as a careful consideration of possible context-dependent effects, may provide a more complete picture of their genetic architecture. Such heterogenous genetic effects may contribute to estimated allelic effects that are infinitesimally small (and difficult to detect) within the population as a whole, even in very large samples. Given smoking’s heavy burden on human health, there is a strong incentive to continue to pursue evidence of heterogeneous effects that can disproportionally burden certain groups.

## Materials and Methods

### Genotypes, phenotypes and moderators

All genotypes, phenotypes, and moderators were obtained from the UK Biobank[23]. Phenotype definitions for smoking behaviors matched exactly GSCAN[11] definitions: Smoking initiation (SI) was a binary phenotype that compared individuals who had smoked at least 100 cigarettes to individuals who had never smoked. Smoking cessation (SC) was a binary phenotype that compared current smokers to former smokers (fields 1239 and 1249). Cigarettes per day (CPD) was based on “Number of cigarettes currently smoked daily (current cigarette smokers)”, “Number of cigarettes previously smoked daily”, or “Number of cigarettes previously smoked daily (current cigar/pipe smokers)” (UK Biobank data fields 2887, 3456, and 6183). Different transformations of CPD were considered, raw CPD, natural log transformed CPD, binned CPD (matching GSCAN defined CPD) included five bins [1 – individuals who smoke(d) 1 to 5; 2 – individuals who smoke(d) 6 to 15; 3 – individuals who smoke(d) 16 to 25; 4 – individuals who smoke(d) 26 to 35; 5 – individuals who smoke(d) 36 to 140], and dichotomized CPD (individuals who smoke more than 20 cigarettes per day vs individuals who smoke 10 or less, excluding remaining individuals). Age of smoking initiation (ASI) was defined as the age at which an individual began smoking regularly (fields 3426 and 2867). Data from the UK Biobank mental health questionnaire was used to construct MDD DSMV-like, and GAD DSMV-like records. GAD DSMV-like cases required endorsement of either Field IDs 20425 or 20542, and endorsement of 20421 with 20420 reported as ≥56 months, and endorsement of 20540 or 20543≥2, and endorsement of 20541 or 20537 or 20539, as well as three or more “Yes” responses to the following symptom Field IDs: 20426 or 20423, 20429, 20419, 20422, 20417, 20427, and endorsement of ‘a little’ or more of field 20418 (impairment or impact). Individuals with complete data but who did not meet the above criteria were treated as controls. Supplementary Figure S89 visually describes the assignment of GAD DSMV-like cases and controls. Similarly, MDD DSMV-like cases required “Yes” responses to 5 or more of the following symptom Field IDs: 20446, 20441, 20533, 20534, 20535, 20449, 20536, 20450, 20435, and 20437, as well as “somewhat” or more response to field 20440, a “almost every day” or more response to field 20439, and a “about half of the day” or more response to 20436.

### Estimating within-strata variance components and between-strata genetic correlations

Using GCTA’s[25] bivariate model implementation (https://cnsgenomics.com/software/gcta/#BivariateGREMLanalysis), we fit a bivariate model treating the same trait—measured in different strata—as two different phenotypes. For example, we modeled the genetic (co)variance of CPD measured in MDD-like cases and CPD measured in MDD-like controls. For a particular trait-by-moderator combination, we built a genetic relationship matrix (GRM) using filtered, genotyped SNPs. Filtering of SNPs was performed using 436,065 European-ancestry individuals after removing individuals with mismatched self-reported and genetic sex, |F_het_|≥0.2, and/or no phenotypic information, where SNPs were removed if they had a genotyping rate or MAF less than 0.05 or had a *p-*value from a Hardy-Weinberg test smaller than 1×10^-8^. GRM entries were then pruned using a relatedness cutoff of 0.05. Fixed effect covariates consisted of sex, batch, assessment center, education level, Townsend deprivation index, age, age squared, and the first 10 genomic principle components, derived from both the European subset of the UK Biobank and the whole UK biobank. When analyzing CPD, current vs former smoker status was included as an additional covariate. Point estimates and standard errors of within-strata variance components and heritabilities were obtained from GCTA, as were between-strata genetic covariances and correlations. Standard errors of estimated variance component fold differences (e.g 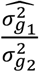, where 1 and 2 index opposing strata) were approximated using the delta method[39], utilizing the sampling variances/covariances of model parameters obtained from GCTA. Specifically, the variance of a ratio of genetic variance estimates was approximated by 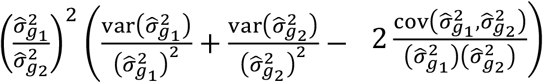, with the ratio of residual variances between strata approximated similarly. The genetic correlation is defined as 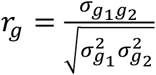 and testing *H*_0_: *r*_*g*_ = 1 was done using a likelihood ratio test, comparing the full model (above) to one in which *r*_*g*_ is constrained to one.

### Modeling the proportion of variance explained by heterogeneous effects

GCTA’s univariate model[25], using the –gxe argument (https://cnsgenomics.com/software/gcta/#GREMLanalysis) was used to decompose the total variance across strata into a shared component 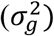, a deviation from the shared component 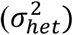, and a residual component 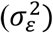 using the same GRM and fixed effects covariates as before. Using GCTAs model parameter estimates, we estimated the proportion of phenotypic variance explained by heterogeneous effects, 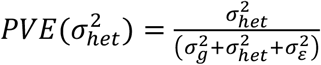, with the standard error provided by GCTA, and tested 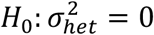 using a likelihood ratio test, comparing the likelihood of the full model to one in which 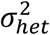 is constrained to 0. For binary traits (dichotomized CPD, SI, and SC), we transformed estimates of 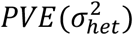 to the liability scale[40], using sample prevalences.

### GWAS, heterogeneous effect inference, and novel SI-loci detection

Stratified GWAS was performed using BOLT-LMM[26]. For all GWAS we used all individuals of European descent, including related individuals. For each BOLT-LMM model fit, fixed effect covariates were identical to those used in GREML based models. GWAS was performed using imputed SNPs with a 0.9 INFO score and 0.01 MAF cutoff, resulting in 7,749,105 SNPs with which to obtain within-strata estimated SNP effects and their standard errors. A z-score was used to infer differences in SNP effects between strata 1 and 2: 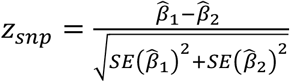, with a two-sided *p*-value: *p*-diff = 2Φ (–|*z*_*snp*_|), where Φ is the normal cumulative distribution function. For all BOLT-LMM model fits, the random polygenic component was estimated from the infinitesimal model as opposed to BOLT’s mixture model. When performing sex-combined GWAS for SI, we combined males and females and ran BOLT similarly while including sex as a covariate. Genomic risk loci and lead SNP identification was performed using FUMA[41] with default parameters. To identify potentially novel SI-associations, we performed sex-combined GWAS using the UK Biobank, then used FUMA to identify genomic risk loci and lead SNPs. We then determined whether a lead SNPs identified from sex-stratified GWAS (associating with males, females, or both) was within an identified sex-combined genomic risk locus. Similarly, we determined whether lead SNPs from the sex-stratified GWAS were within risk loci reported in Liu et al. 2019[11].

### Partitioning stratified heritability estimates to functional categories

Using stratified GWAS results obtained from BOLT-LMM (see above), we performed cell-type specific LDSC analysis[28] to partition strata-specific SNP heritability along functional annotations, *i*.*e*., SNPs within 100kb of genes uniquely expressed in a particular tissue. This was done using within-annotation LD scores, with annotations derived from the baseline model[27], the Cahoy et al. gene-expression dataset[42], GTEx (both multi-tissue assessment of cell-type specific genes and brain-specific assessment of cell-type specific genes)[43], and the Franke Lab gene expression dataset[44,45]. LD scores were downloaded and analysis was carried out according to the steps outlined at https://github.com/bulik/ldsc/wiki/Cell-type-specific-analyses. Only hapmap3 SNPs were used in all LDSC analyses. To infer differences in partitioned heritabilities between strata, firstly we fit the partitioned LDSC model separately for each strata, thus obtaining strata-specific partitioned LDSC estimated coefficients (representing the per-SNP contribution to heritability from a particular annotation) and estimated heritability enrichments (heritability of an annotation divided by the number of SNPs in the annotation). We then inferred differences in LDSC estimated coefficients and heritability enrichments between strata, using the same z-score and two-sided testing approach outlined in the previous section.

### Simulations

All simulations utilized a single causal variant model, whereby genotypes at the causal variant *x* were sampled from the binomial distribution: *x* ∼ *B*(2, *p*), where *p* is the minor allele frequency. Simulating a sex-specific binary trait *y* was done by sampling: *y* ∼ *B*(1, *e*^*xβ*^/1 + *e*^*xβ*^)), with *β* being the sex-specific log odds ratio of the causal variant. To transform linear coefficients *b* (like those obtained from BOLT-LMM) to log odds ratios, we used the approximation 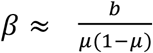, with *μ* being the sex-specific case fraction. For each simulation, sex-specific causal variant effects were fixed and genotypes at the causal variant were randomly sampled for 5000 replicates.

### Polygenic risk scoring and prediction accuracy

To compute SNP effects used in polygenic risk scoring, we used marginal SNP effects from BOLT-LMM model fits, then obtained best linear unbiased predictions (BLUPs) of SNP effects (for all 7,749,105 SNPs) that account for LD using SBLUP [25,46]. In both CADD and Add Health datasets, we used randomly sampled, unrelated individuals of European descent. In each dataset, PRS were computed from SNPs imputed from haplotype reference consortium data (MAF > 0.01, INFO R^2^ > 0.95). In CADD, we predicted the response to “Have you smoked at least 20 cigarettes in your lifetime?” and in Add Health we predicted the response to “Have you ever smoked cigarettes regularly, that is, at least 1 cigarette every day for 30 days?”. To assess prediction accuracy, we compared a full model including PRS and covariates to a reduced model including covariates only. For both datasets, covariates were age, age-squared, sex, educational attainment (coded categorically), and the first 10 genomic principal components.

## Data Availability

Summary stats for all within-strata GWASes and between-strata difference tests will be made available upon publication. Links will be available on FUMA (https://fuma.ctglab.nl/browse).

## Acknowledgements

This work was supported by NIMH Training Grant 5T32MH016880-38, R01 AG046938-06 (MPIs: Reynolds, Wadsworth), R01 DA044283-01 (Vrieze), R01 MH100141-06 (Keller), and awards DA042755 and DA032555 (Hopfer). CADD and GADD data were supported by DA011015, DA035804, DA012845, DA035804, and DA021692. This work utilized the Summit supercomputer, which is supported by the National Science Foundation (awards ACI-1532235 and ACI-1532236), the University of Colorado Boulder, and Colorado State University. The Summit supercomputer is a joint effort of the University of Colorado Boulder and Colorado State University. Data storage supported by the University of Colorado Boulder “PetaLibrary”. This research has been conducted using the UK Biobank Resource (application number 1665). We thank the UK Biobank and the participants of the UK Biobank, and the participants of the AddHealth (PI: Harris) and CADD (PI: Hewitt) studies. Lastly, we thank Robin Corley for valuable contributions to the writing of this manuscript.

