## Supplementary Note for "Investigating heterogeneous genetic effects contributing to smoking behaviors"

Investigating heterogeneous genetic effects contributing to smoking behaviors: Supplementary Note

### Discussion

#### Novel SI-associated loci identified through sex-stratified GWAS

We identified 6 GWS loci using sex-stratified GWAS for SI that were not detected in a sex-combined GWAS of the UK Biobank, nor detected in the largest known GWAS meta-analysis of SI[1]. The first of these loci, marked by rs6547148, is within an intron of *LRRTM4*, a gene predominantly expressed in pituitary, nerve, and numerous brain tissues[2]. This is a gene containing variants previously associated with insomnia ($p$-value = $3x10^{-9}$)[3], schizophrenia ($p$-value = $1x10^{-7}$)[4], and forced vital capacity ($p$-value = $1x10^{-6}$) [5]. Another novel signal, rs360892, is a known splicing QTL for *IQSEC1* expression in the testis ($p$-value = $3.8x10^{-21}$) [2], offering one possible explanation for sex differences in genetic mechanisms. *IQSEC1* is also known to contain variants associated with BMI ($p$-value = $2x10^{-10}$)[6]. rs7770532 is within 10Kb of SNPs associated with medication use pertaining to the renin-angiotensin system (RAS) ($p$-value = $1x10^{-8}$)[7], which is intriguing given that RAS homeostasis is known to be affected by nicotine[8]. Likewise, rs76759272 is within 10Kb of SNPs associated with eosinophil counts ($p$-value = $8x10^{-9}$)[9], which are known to be affected by smoking[10]. rs669257, not identified by Liu et al.[1] using a univariate GWAS for SI, is an eQTL for *MIR4697HG* ($p$-value = $1.5x10^{-24}$) and *IGSF9B* ($p$-value = $7.5x10^{-13}$) in the tibial nerve [2], and was identified by a multivariate GWAS of smoking and alcohol traits using MTAG[11]. Likewise, rs11066972 is within 1Kb of a SNP associated with smoking status (ever vs never smokers)[12], a different definition of smoking initiation from what was used in this study.
