## Supplementary Figures for "Investigating heterogeneous genetic effects contributing to smoking behaviors"

### Investigating heterogeneous genetic effects contributing to smoking behaviors: Supplementary Figures

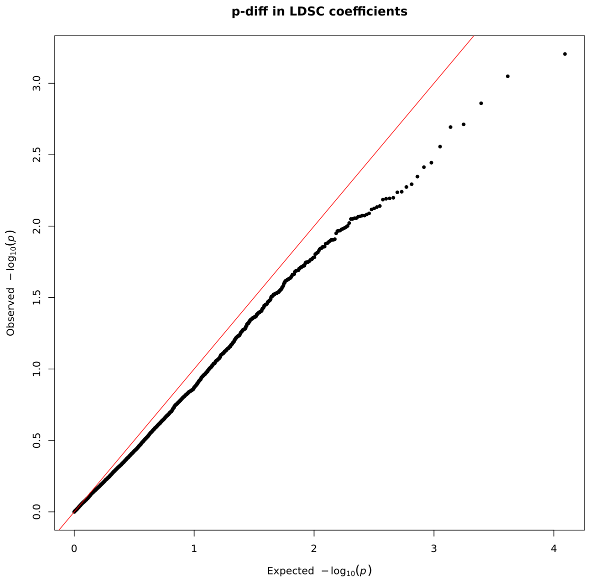

Figure S1: QQ plot showing the distribution of p-diff when testing for differences in partitioned LDSC coefficients between strata. All trait-by-moderator combinations are shown.

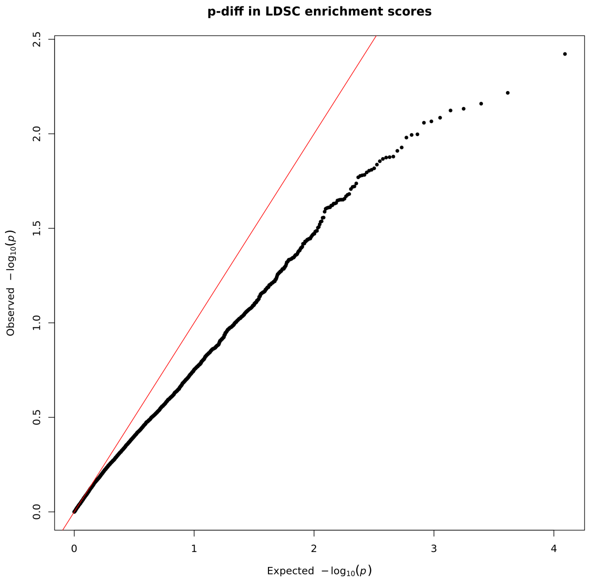

Figure S2: QQ plot showing the distribution of p-diff when testing for differences in partitioned LDSC enrichment scores between strata. All trait-by-moderator combinations are shown.

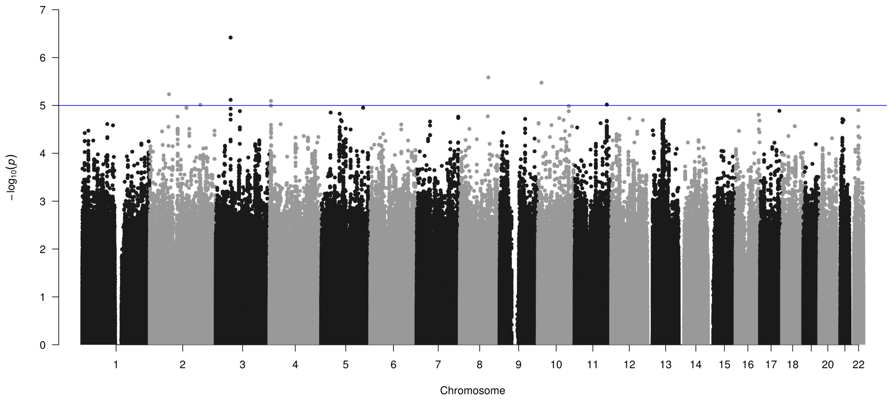

Figure S3: Testing differences in CPD (binned) SNP effects between late-onset and early-onset smokers. Blue line indicates $p$-diff $< 1\text{x}10^{-5}$. Red line indicates $p$-diff $< 5\text{x}10^{-8}$.

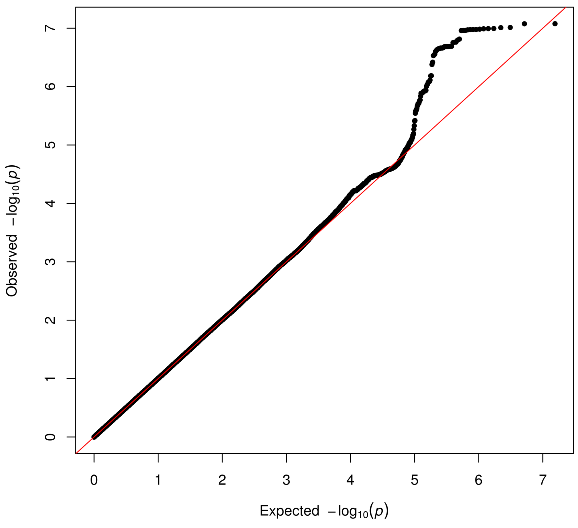

Figure S4: QQ plot showing the distribution of $p$-diff when testing for differences in CPD (binned) SNP effects between late-onset and early-onset smokers.

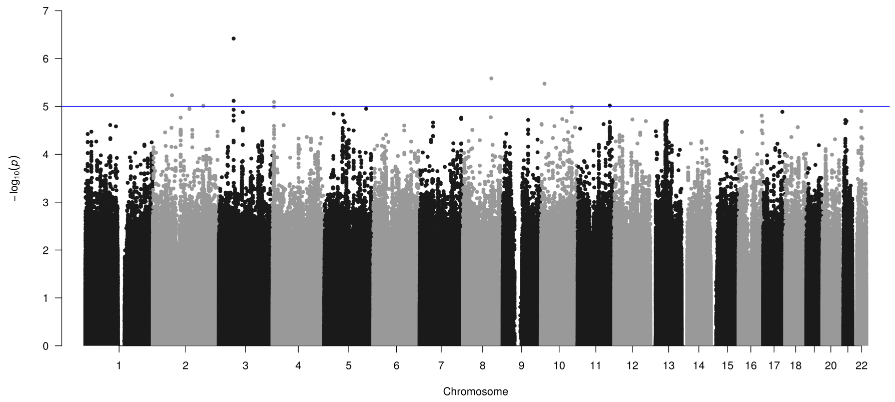

Figure S5: Testing differences in CPD (binned) SNP effects between GAD cases and controls. Blue line indicates $p$-diff < 1x10$^{-5}$. Red line indicates $p$-diff < 5x10$^{-8}$.

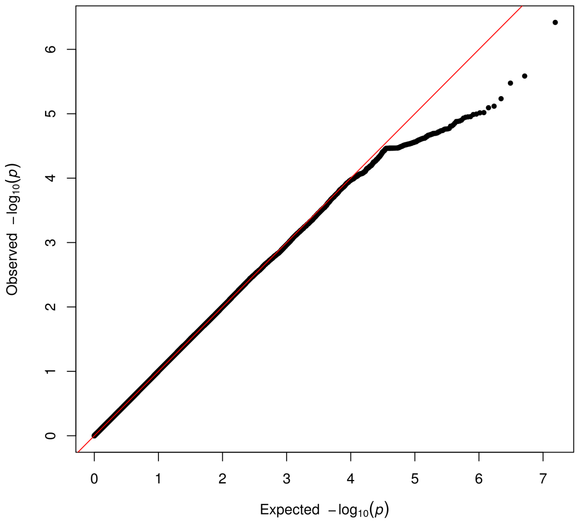

Figure S6: QQ plot showing the distribution of $p$-diff when testing for differences in CPD (binned) SNP effects between GAD cases and controls.

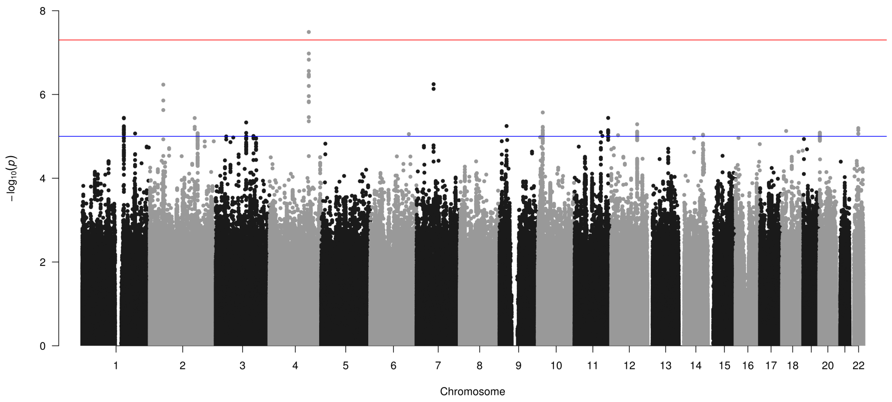

Figure S7: Testing differences in CPD (binned) SNP effects between MDD cases and controls. Blue line indicates $p$-diff < 1x10$^{-5}$. Red line indicates $p$-diff < 5x10$^{-8}$.

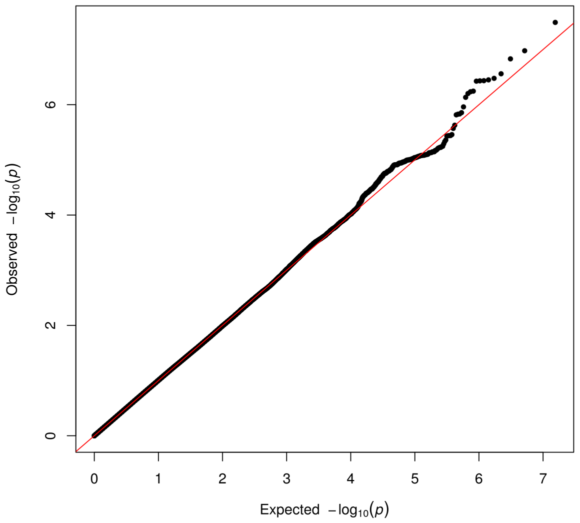

Figure S8: QQ plot showing the distribution of $p$-diff when testing for differences in CPD (binned) SNP effects between MDD cases and controls.

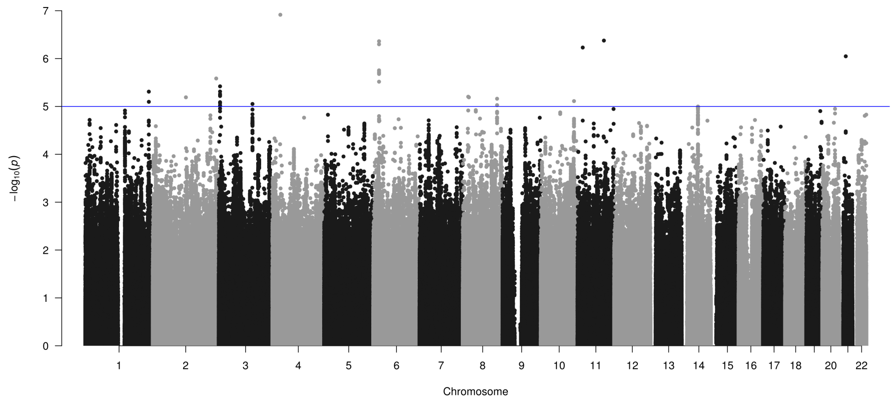

Figure S9: Testing differences in CPD (binned) SNP effects between PSYCH cases and controls. Blue line indicates $p$-diff $< 1x10^{-5}$. Red line indicates $p$-diff $< 5x10^{-8}$.

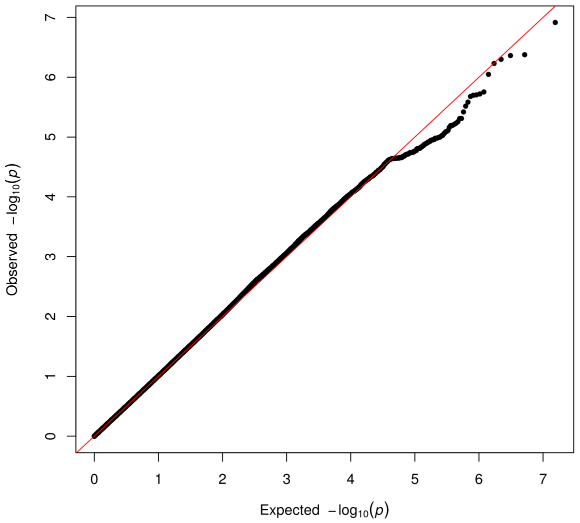

Figure S10: QQ plot showing the distribution of $p$-diff when testing for differences in CPD (binned) SNP effects between PSYCH cases and controls.

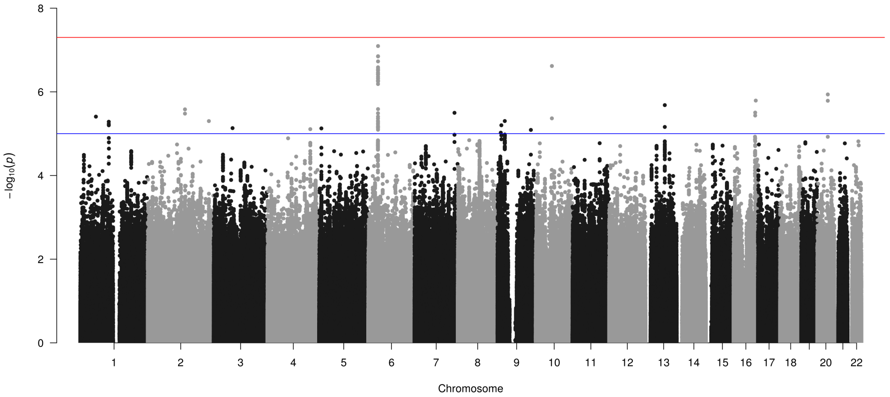

Figure S11: Testing differences in CPD (binned) SNP effects between sexes. Blue line indicates $p$-diff $<$ 1x10$^{-5}$. Red line indicates $p$-diff $<$ 5x10$^{-8}$.

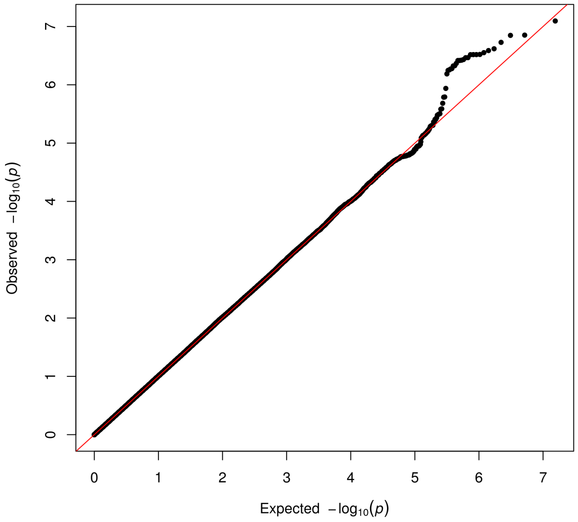

Figure S12: QQ plot showing the distribution of $p$-diff when testing for differences in CPD (binned) SNP effects between sexes.

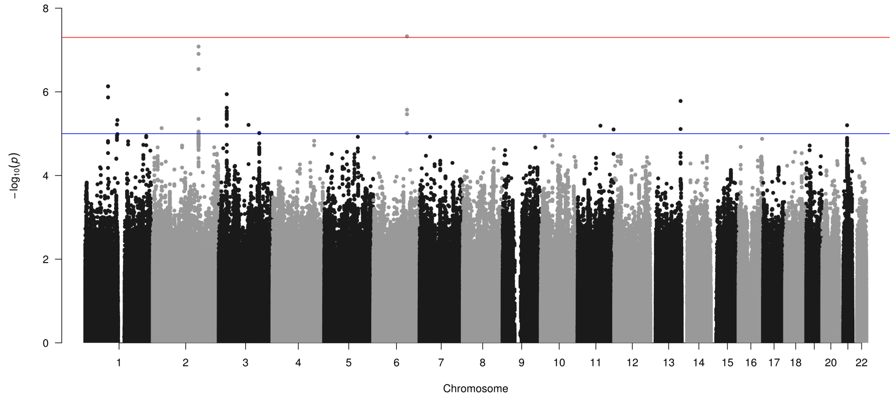

Figure S13: Testing differences in CPD (dichotomized) SNP effects between late-onset and early-onset smokers. Blue line indicates $p$-diff $< 1x10^{-5}$. Red line indicates $p$-diff $< 5x10^{-8}$.

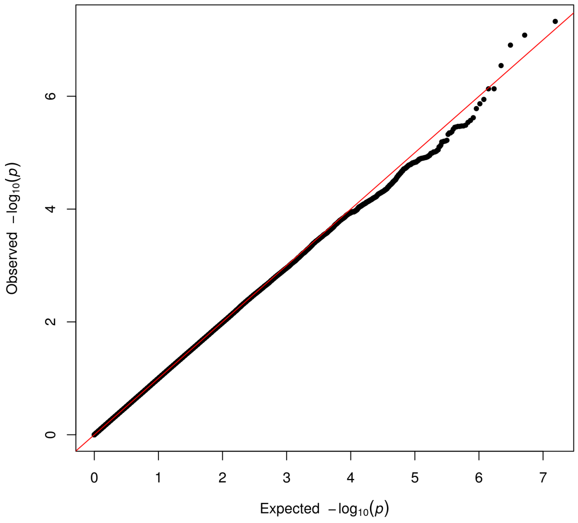

Figure S14: QQ plot showing the distribution of $p$-diff when testing for differences in CPD (dichotomized) SNP effects between late-onset and early-onset smokers.

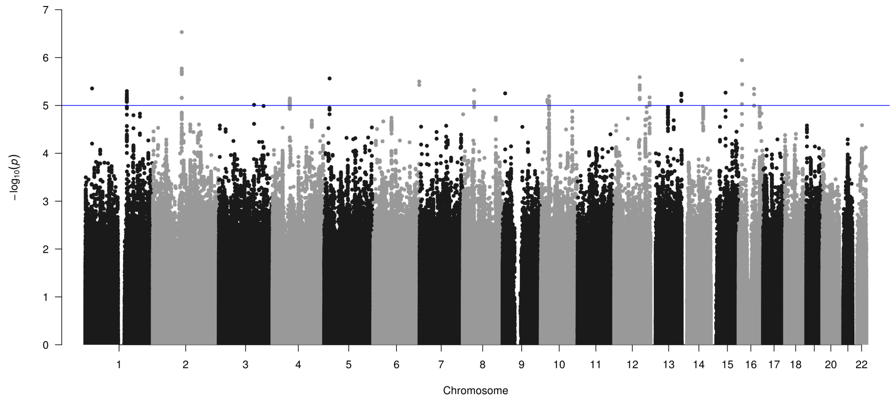

Figure S15: Testing differences in CPD (dichotomized) SNP effects between MDD cases and controls. Blue line indicates $p$-diff < 1x10$^{-5}$. Red line indicates $p$-diff < 5x10$^{-8}$.

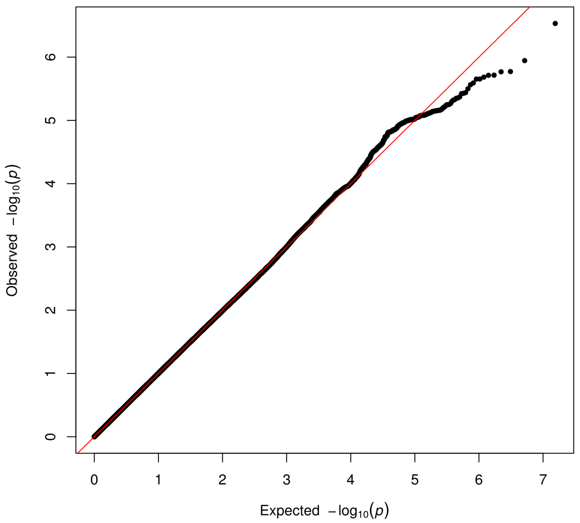

Figure S16: QQ plot showing the distribution of $p$-diff when testing for differences in CPD (dichotomized) SNP effects between MDD cases and controls.

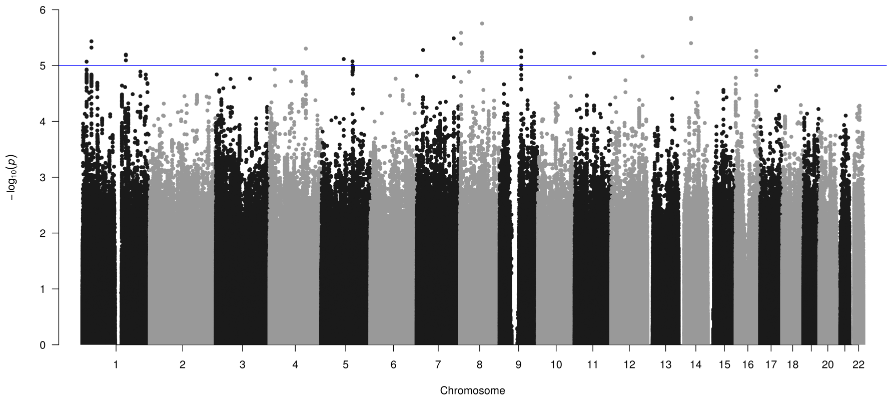

Figure S17: Testing differences in CPD (dichotomized) SNP effects between PSYCH cases and controls. Blue line indicates $p$-diff $< 1\text{x}10^{-5}$. Red line indicates $p$-diff $< 5\text{x}10^{-8}$.

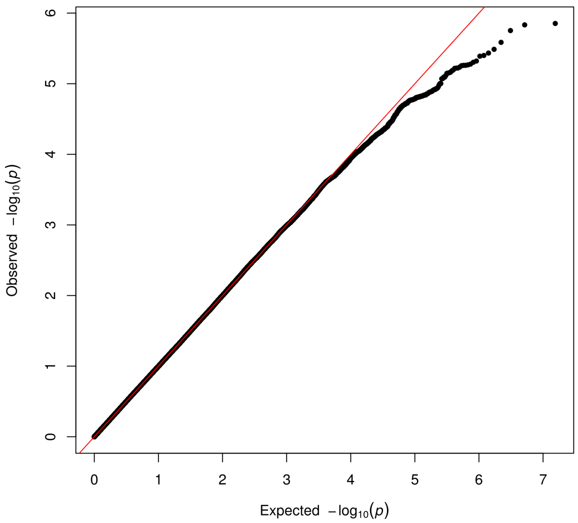

Figure S18: QQ plot showing the distribution of $p$-diff when testing for differences in CPD (dichotomized) SNP effects between PSYCH cases and controls.

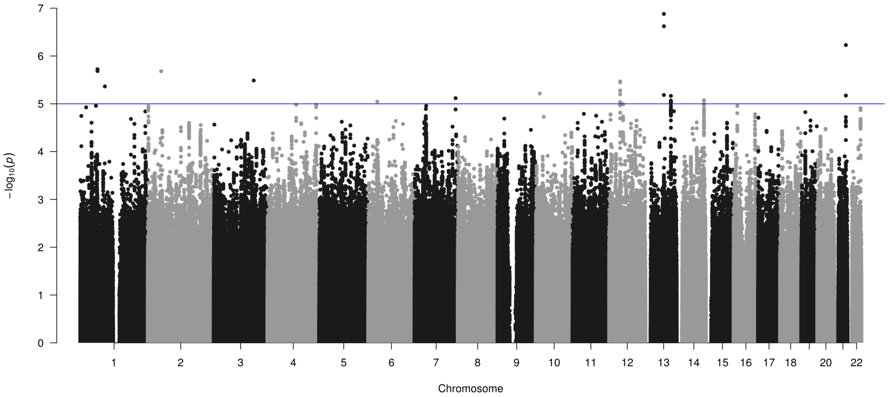

Figure S19: Testing differences in CPD (dichotomized) SNP effects between sexes. Blue line indicates $p$-diff < 1x10$^{-5}$. Red line indicates $p$-diff < 5x10$^{-8}$.

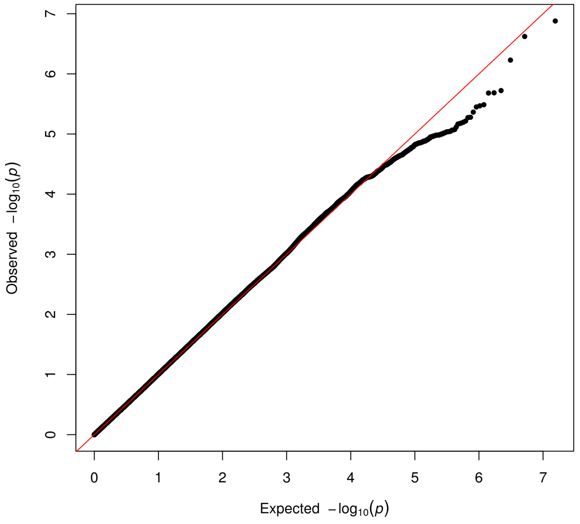

Figure S20: QQ plot showing the distribution of $p$-diff when testing for differences in CPD (dichotomized) SNP effects between sexes.

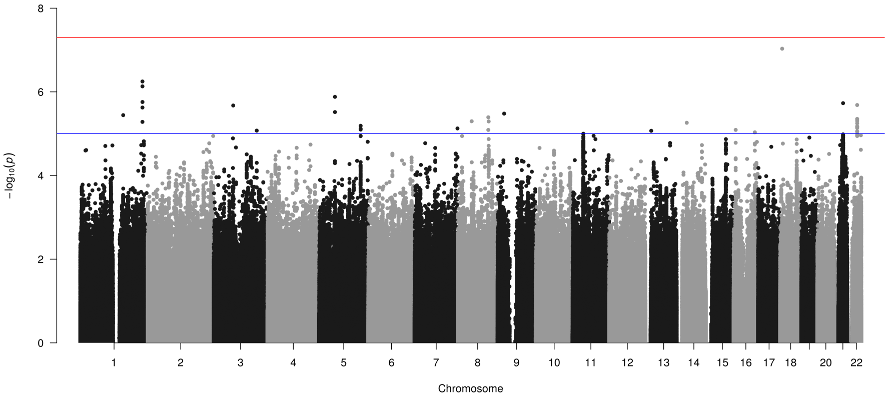

Figure S21: Testing differences in CPD (log transformed) SNP effects between late-onset and early-onset smokers. Blue line indicates $p$-diff $< 1\text{x}10^{-5}$. Red line indicates $p$-diff $< 5\text{x}10^{-8}$.

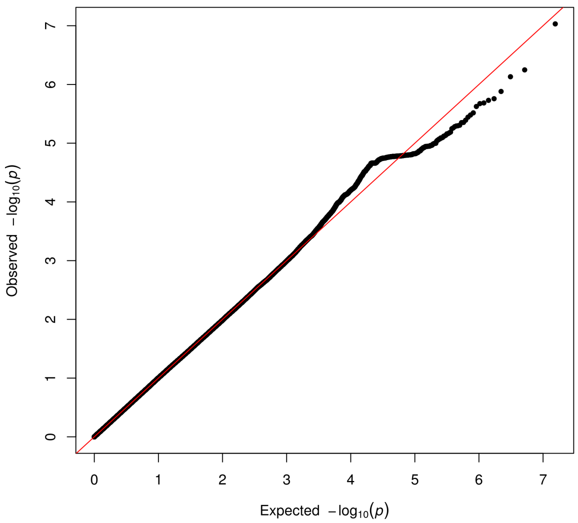

Figure S22: QQ plot showing the distribution of $p$-diff when testing for differences in CPD (log transformed) SNP effects between late-onset and early-onset smokers.

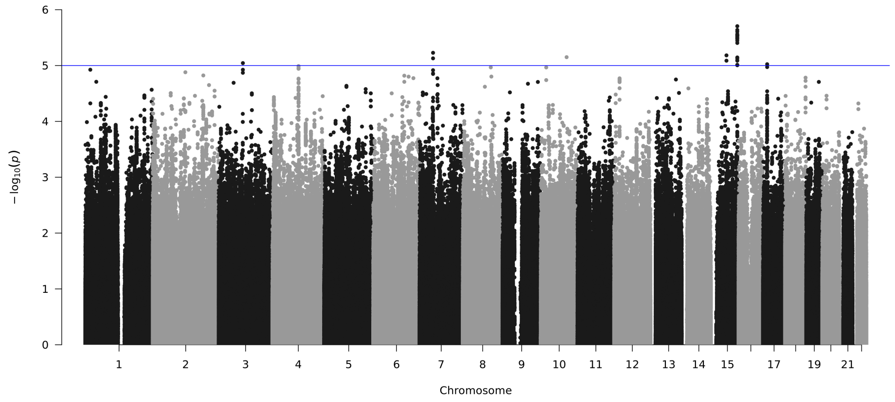

Figure S23: Testing differences in CPD (log transformed) SNP effects between GAD cases and controls. Blue line indicates $p$-diff < 1x10$^{-5}$. Red line indicates $p$-diff < 5x10$^{-8}$.

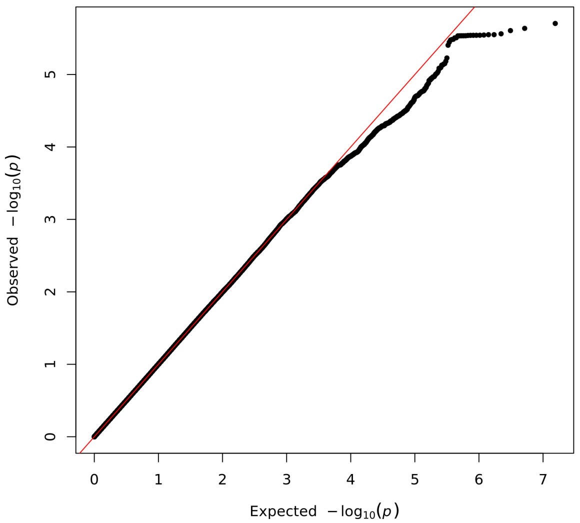

Figure S24: QQ plot showing the distribution of $p$-diff when testing for differences in CPD (log transformed) SNP effects between GAD cases and controls.

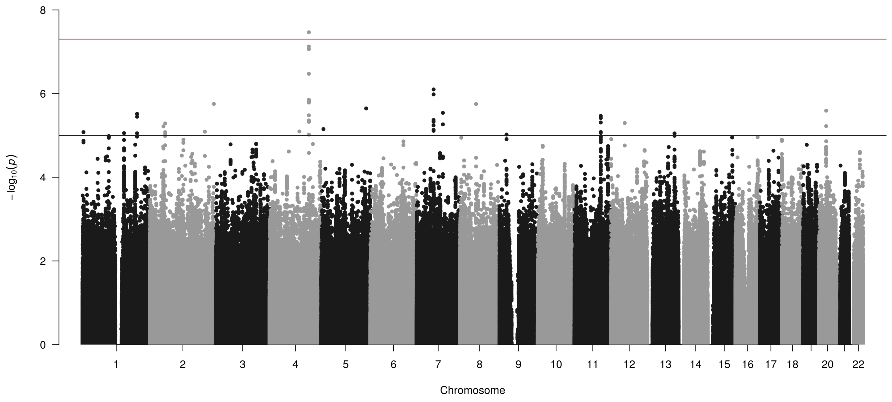

Figure S25: Testing differences in CPD (log transformed) SNP effects between MDD cases and controls. Blue line indicates $p$-diff $< 1\text{x}10^{-5}$. Red line indicates $p$-diff $< 5\text{x}10^{-8}$.

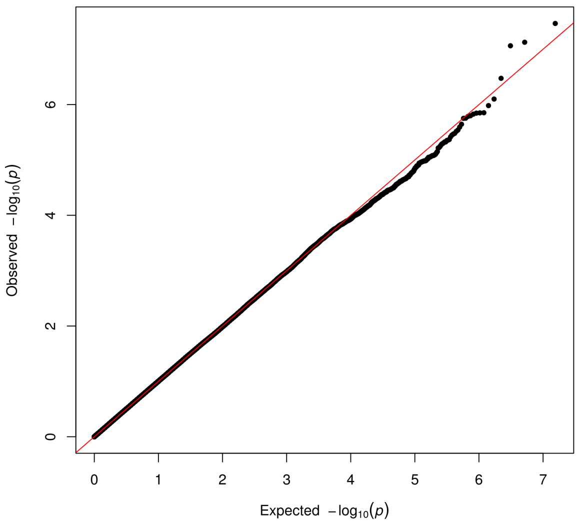

Figure S26: QQ plot showing the distribution of $p$-diff when testing for differences in CPD (log transformed) SNP effects between MDD cases and controls.

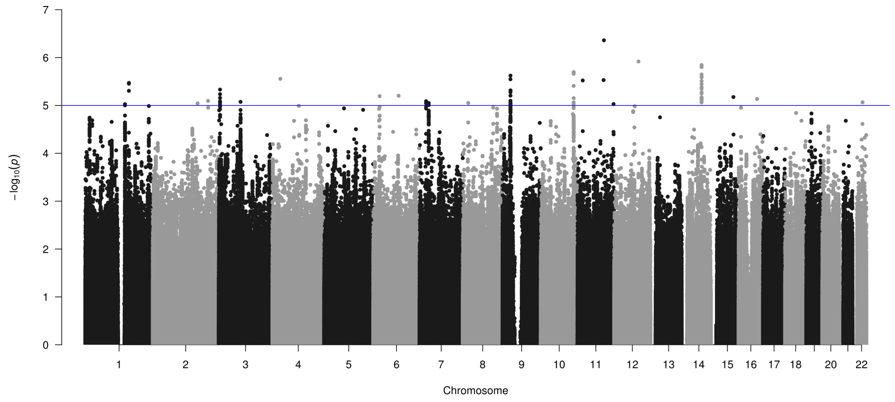

Figure S27: Testing differences in CPD (log transformed) SNP effects between PSYCH cases and controls. Blue line indicates $p$-diff $< 1x10^{-5}$. Red line indicates $p$-diff $< 5x10^{-8}$.

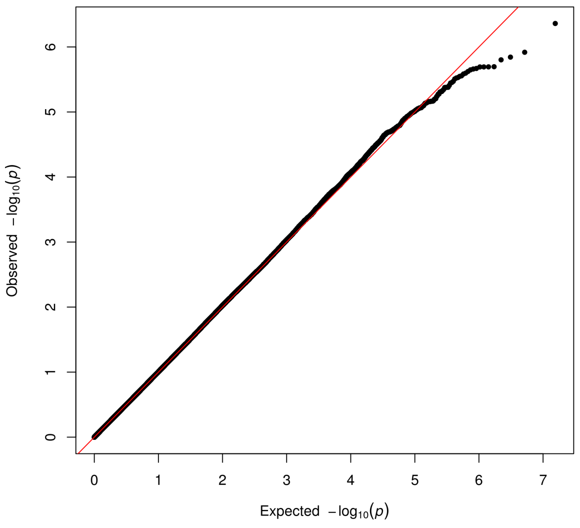

Figure S28: QQ plot showing the distribution of $p$-diff when testing for differences in CPD (log transformed) SNP effects between PSYCH cases and controls.

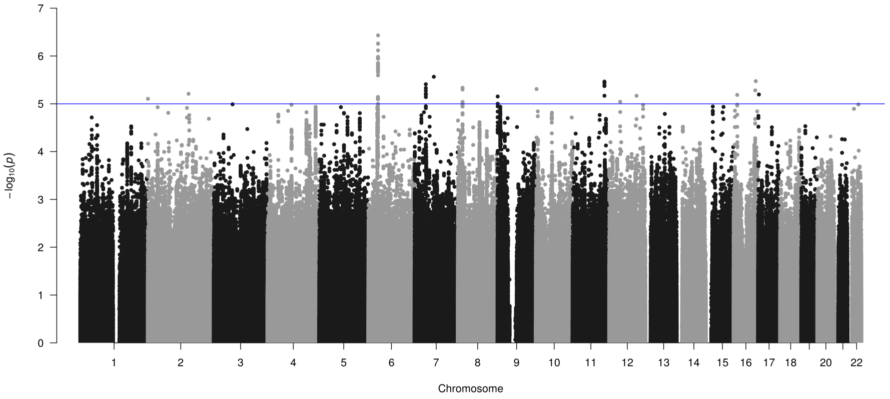

Figure S29: Testing differences in CPD (log transformed) SNP effects between sexes. Blue line indicates $p$-diff < 1x10$^{-5}$. Red line indicates $p$-diff < 5x10$^{-8}$.

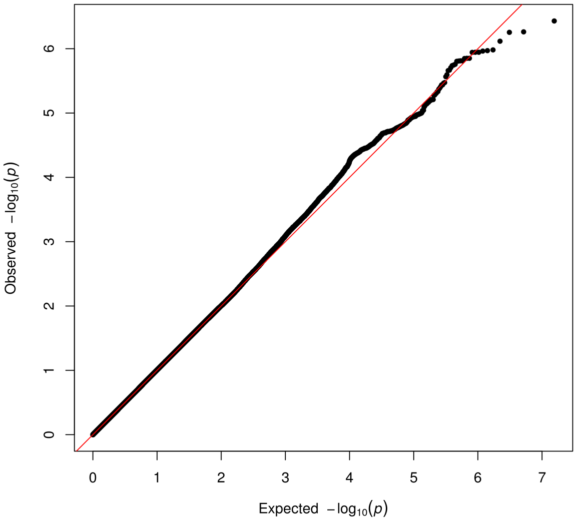

Figure S30: QQ plot showing the distribution of $p$-diff when testing for differences in CPD (log transformed) SNP effects between sexes.

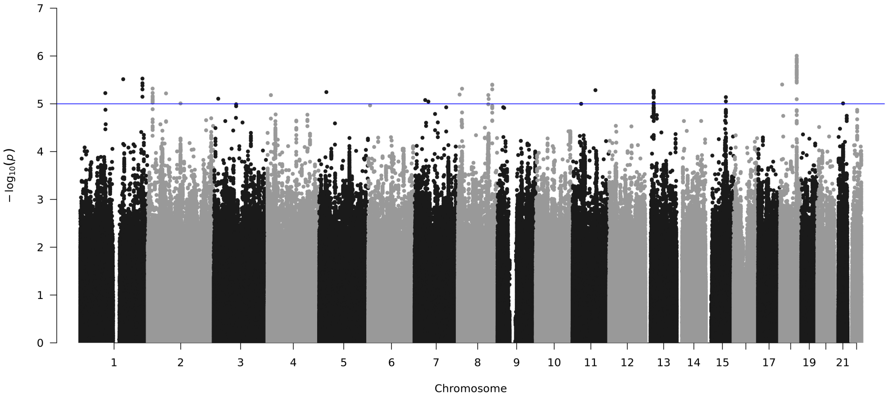

Figure S31: Testing differences in CPD (raw scale) SNP effects between late-onset and early-onset smokers. Blue line indicates $p$-diff $< 1x10^{-5}$. Red line indicates $p$-diff $< 5x10^{-8}$.

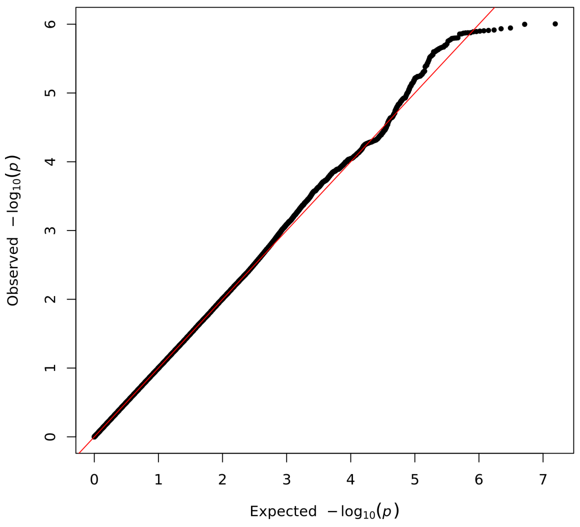

Figure S32: QQ plot showing the distribution of $p$-diff when testing for differences in CPD (raw scale) SNP effects between late-onset and early-onset smokers.

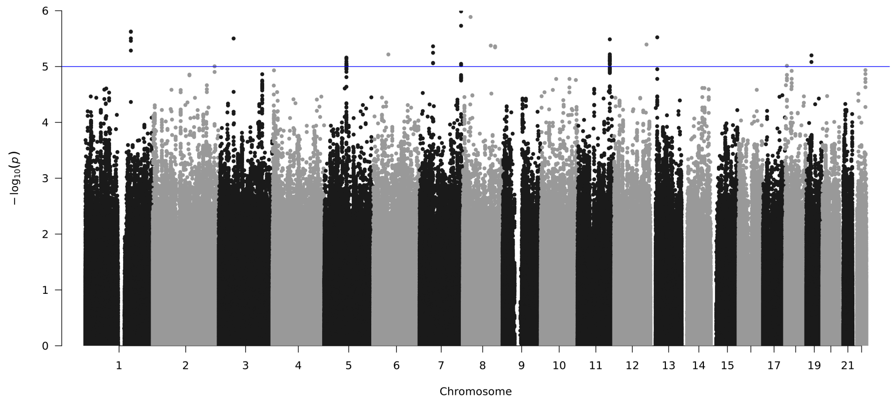

Figure S33: Testing differences in CPD (raw scale) SNP effects between GAD cases and controls. Blue line indicates $p$-diff < 1x10$^{-5}$. Red line indicates $p$-diff < 5x10$^{-8}$.

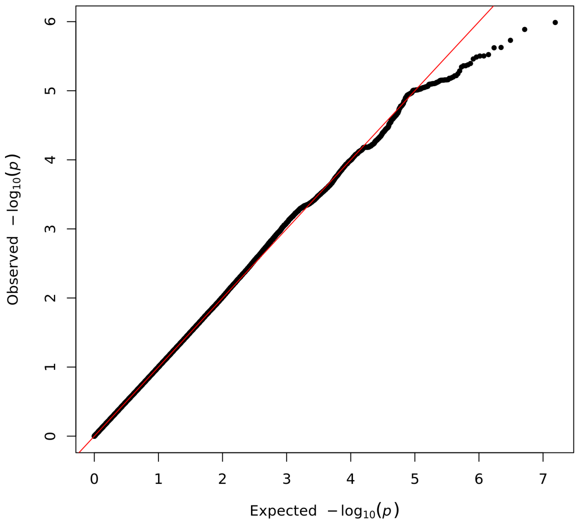

Figure S34: QQ plot showing the distribution of $p$-diff when testing for differences in CPD (raw scale) SNP effects between GAD cases and controls.

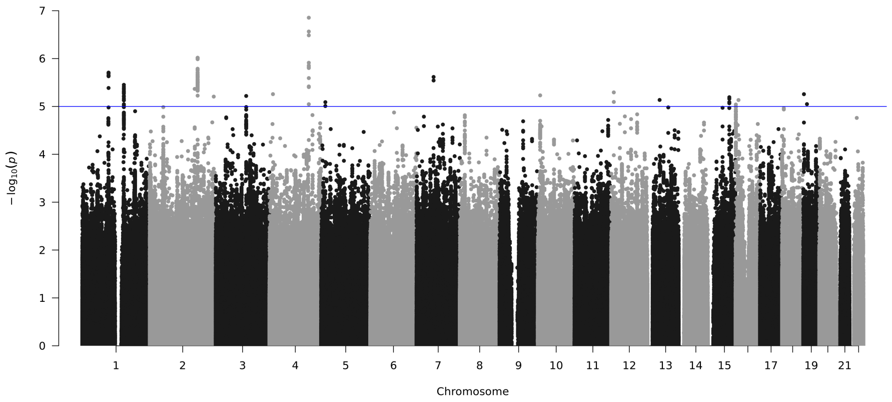

Figure S35: Testing differences in CPD (raw scale) SNP effects between MDD cases and controls. Blue line indicates $p$-diff < 1x10$^{-5}$. Red line indicates $p$-diff < 5x10$^{-8}$.

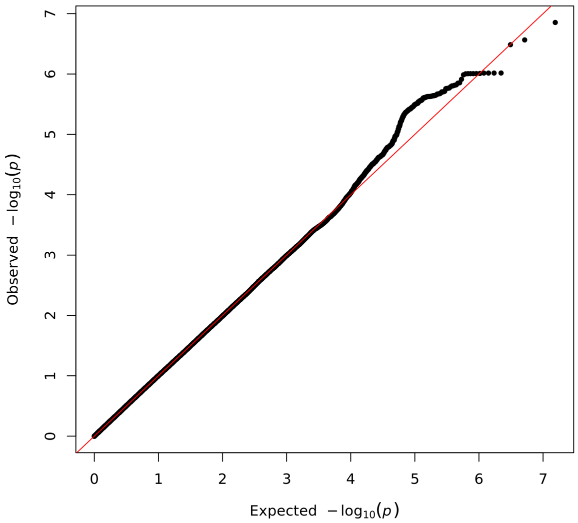

Figure S36: QQ plot showing the distribution of $p$-diff when testing for differences in CPD (raw scale) SNP effects between MDD cases and controls.

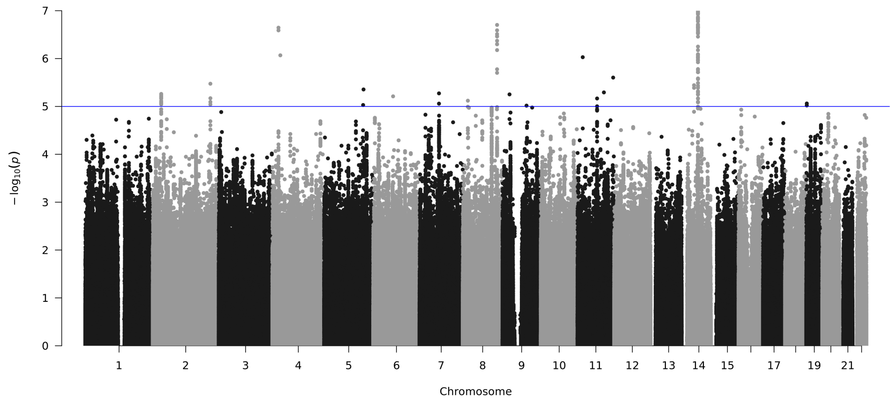

Figure S37: Testing differences in CPD (raw scale) SNP effects between PSYCH cases and controls. Blue line indicates $p$-diff $< 1\text{x}10^{-5}$. Red line indicates $p$-diff $< 5\text{x}10^{-8}$.

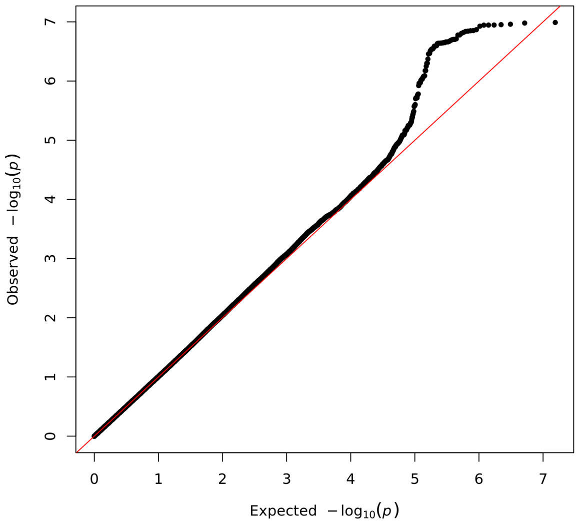

Figure S38: QQ plot showing the distribution of $p$-diff when testing for differences in CPD (raw scale) SNP effects between PSYCH cases and controls.

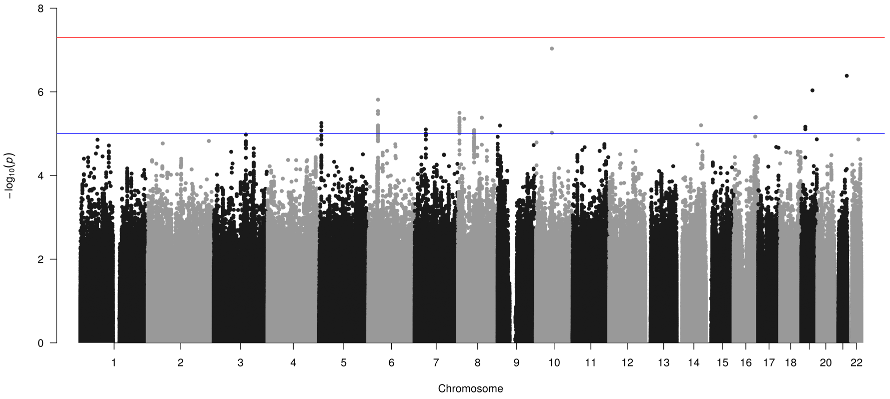

Figure S39: Testing differences in CPD (raw scale) SNP effects between sexes. Blue line indicates $p$-diff $<$ 1x10$^{-5}$. Red line indicates $p$-diff $<$ 5x10$^{-8}$.

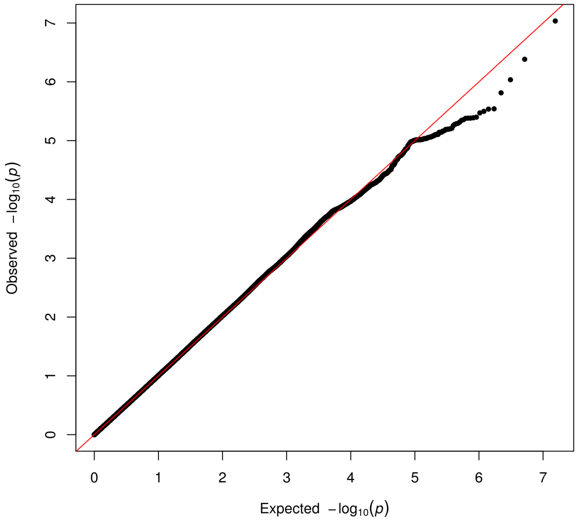

Figure S40: QQ plot showing the distribution of $p$-diff when testing for differences in CPD (raw scale) SNP effects between sexes.

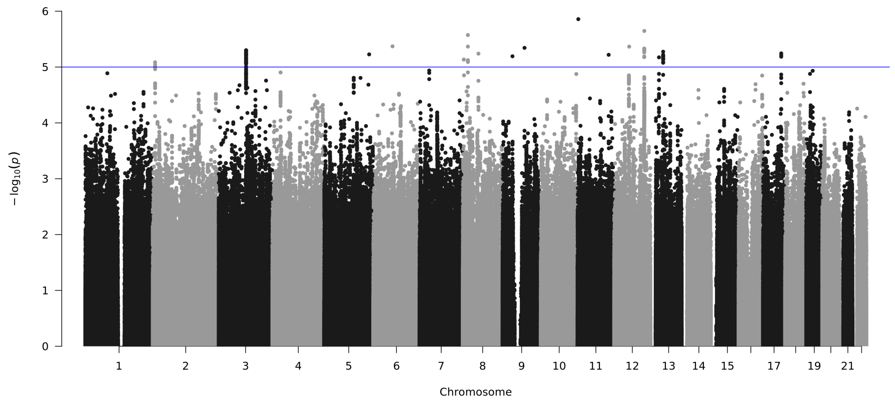

Figure S41: Testing differences in SC SNP effects between late-onset and early-onset smokers. Blue line indicates $p$-diff < 1x10$^{-5}$. Red line indicates $p$-diff < 5x10$^{-8}$.

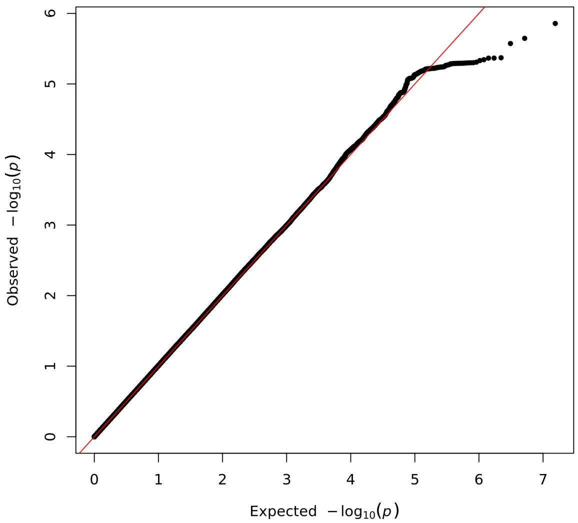

Figure S42: QQ plot showing the distribution of $p$-diff when testing for differences in SC SNP effects between late-onset and early-onset smokers.

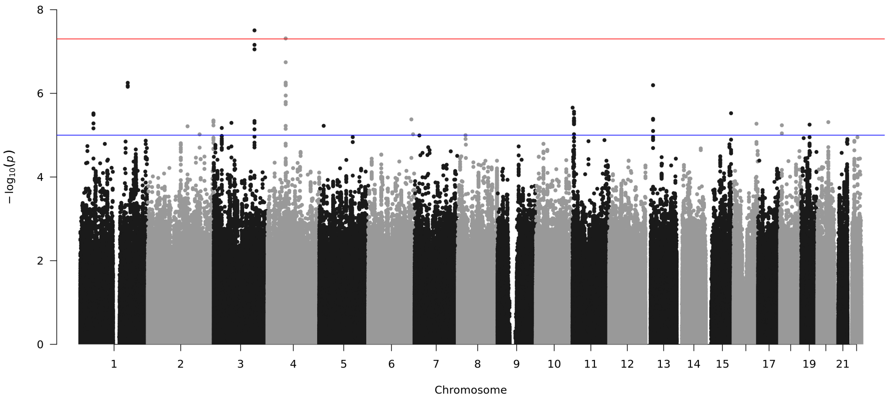

Figure S43: Testing differences in SC SNP effects between GAD cases and controls. Blue line indicates $p$-diff $< 1x10^{-5}$. Red line indicates $p$-diff $< 5x10^{-8}$.

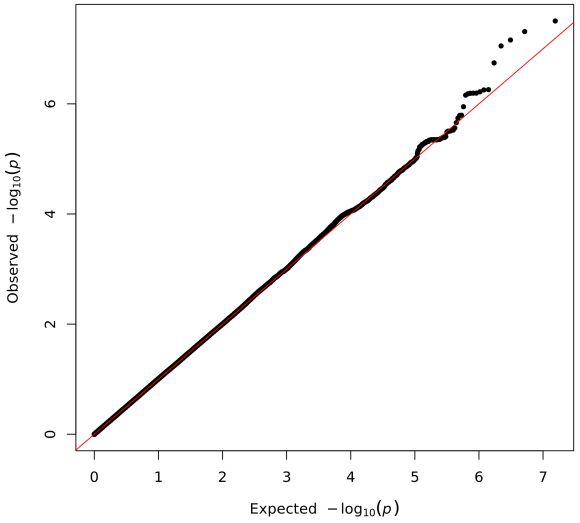

Figure S44: QQ plot showing the distribution of $p$-diff when testing for differences in SC SNP effects between GAD cases and controls.

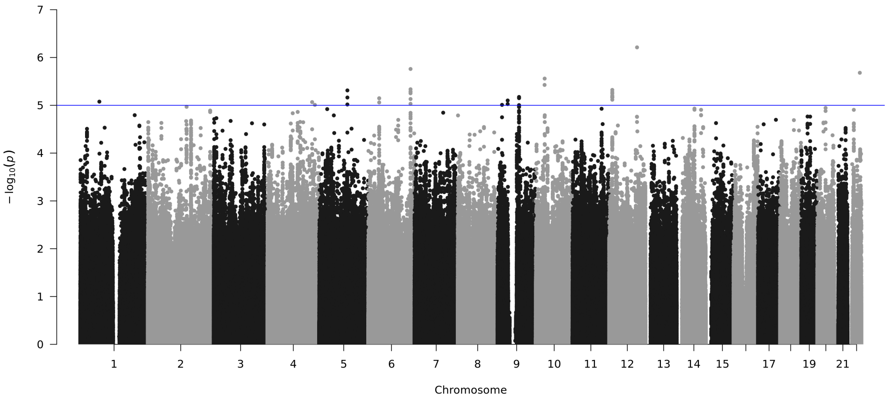

Figure S45: Testing differences in SC SNP effects between MDD cases and controls. Blue line indicates $p$-diff $< 1x10^{-5}$. Red line indicates $p$-diff $< 5x10^{-8}$.

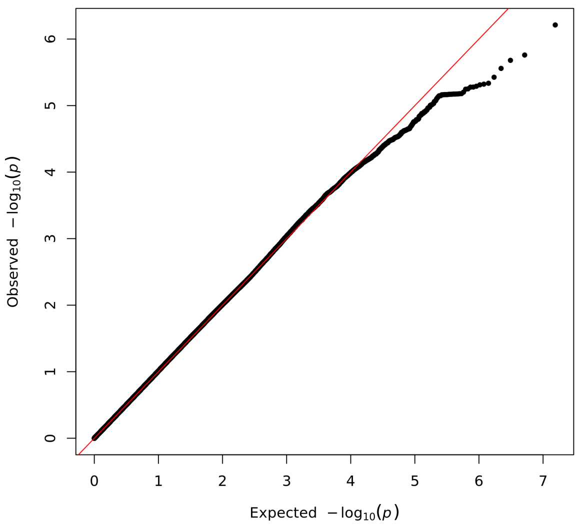

Figure S46: QQ plot showing the distribution of $p$-diff when testing for differences in SC SNP effects between MDD cases and controls.

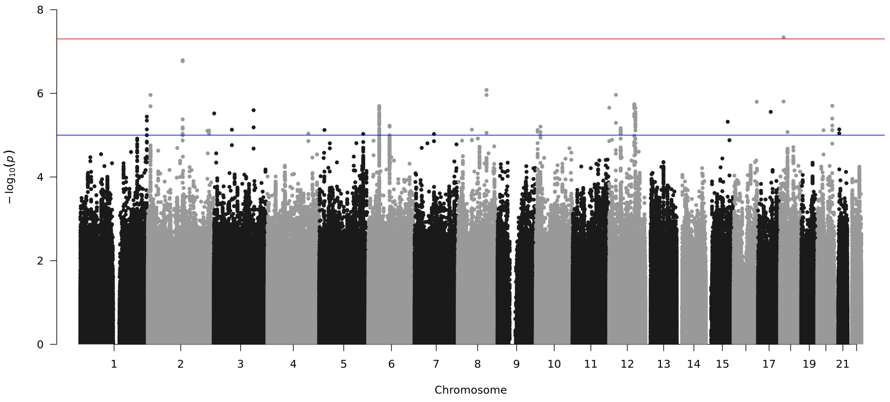

Figure S47: Testing differences in SC SNP effects between PSYCH cases and controls. Blue line indicates $p$-diff $< 1\text{x}10^{-5}$. Red line indicates $p$-diff $< 5\text{x}10^{-8}$.

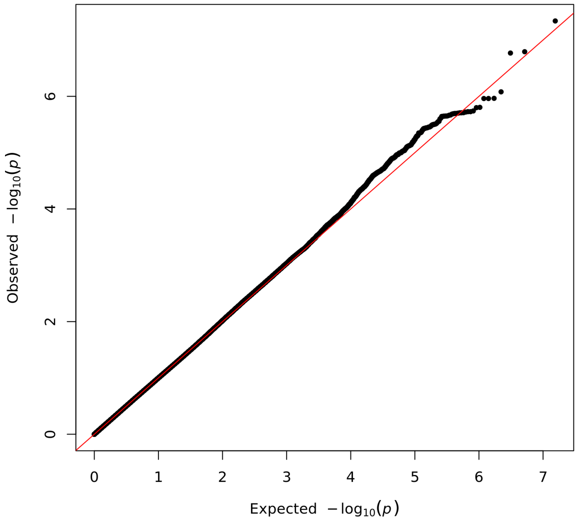

Figure S48: QQ plot showing the distribution of $p$-diff when testing for differences in SC SNP effects between PSYCH cases and controls.

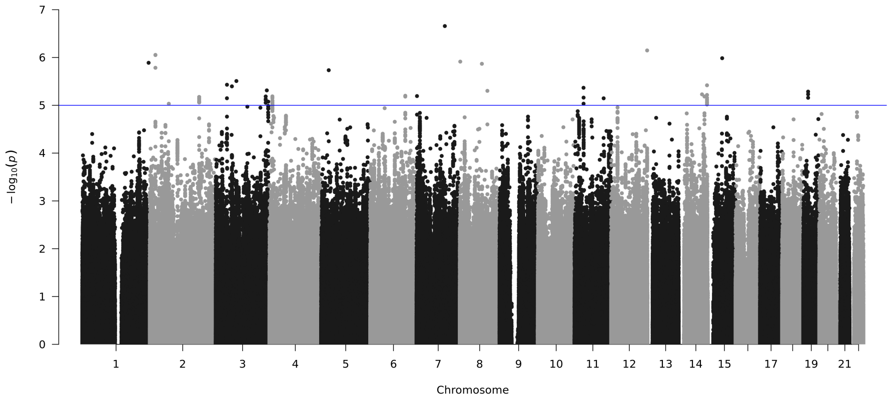

Figure S49: Testing differences in SC SNP effects between sexes. Blue line indicates $p$-diff < 1x10$^{-5}$. Red line indicates $p$-diff < 5x10$^{-8}$.

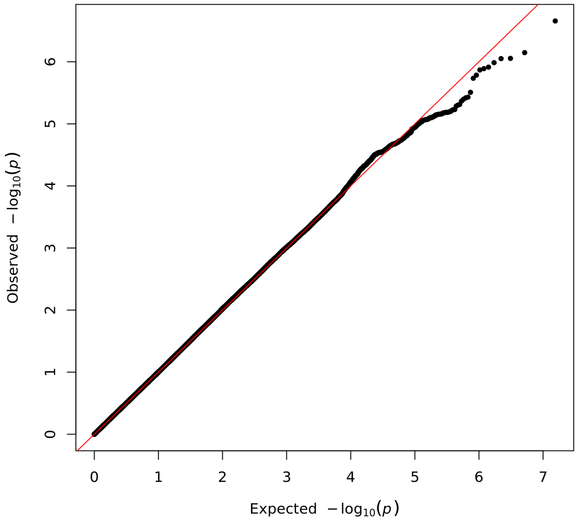

Figure S50: QQ plot showing the distribution of $p$-diff when testing for differences in SC SNP effects between sexes.

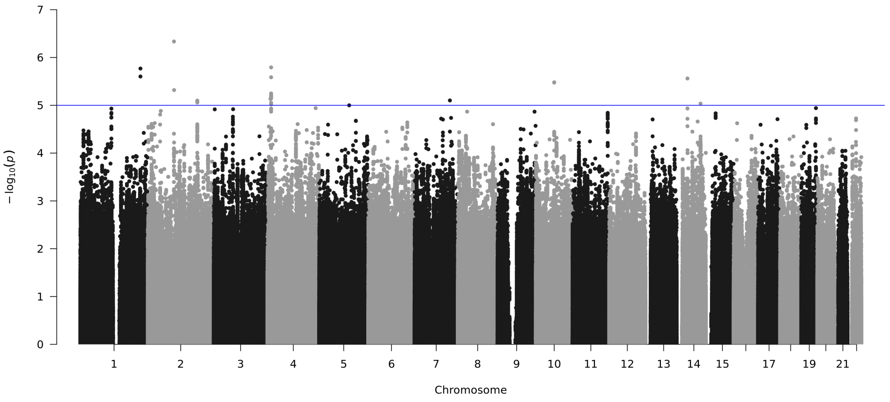

Figure S51: Testing differences in SI SNP effects between GAD cases and controls. Blue line indicates $p$-diff $< 1\text{x}10^{-5}$. Red line indicates $p$-diff $< 5\text{x}10^{-8}$.

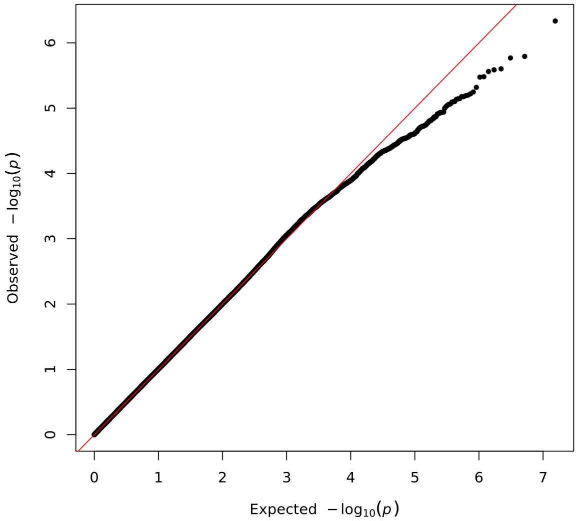

Figure S52: QQ plot showing the distribution of $p$-diff when testing for differences in SI SNP effects between GAD cases and controls.

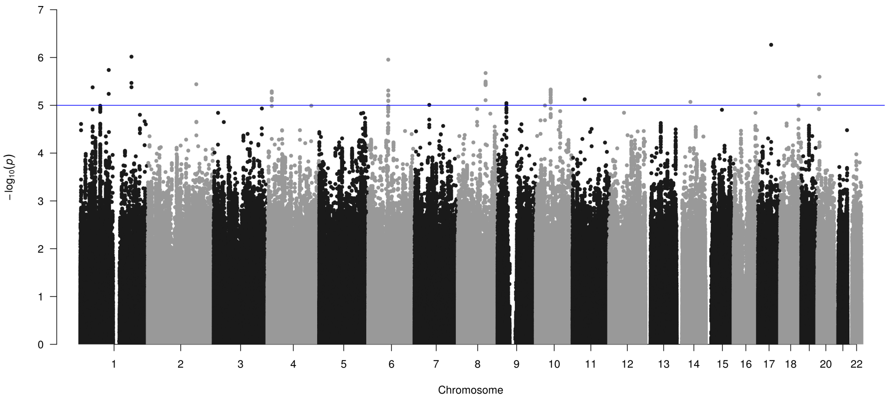

Figure S53: Testing differences in SI SNP effects between MDD cases and controls. Blue line indicates $p$-diff $< 1\text{x}10^{-5}$. Red line indicates $p$-diff $< 5\text{x}10^{-8}$.

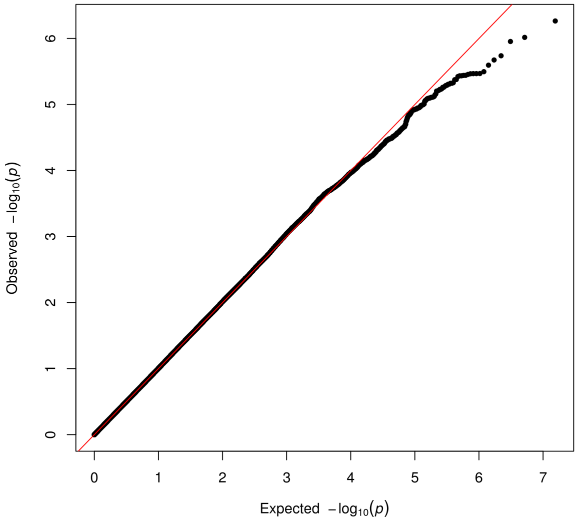

Figure S54: QQ plot showing the distribution of $p$-diff when testing for differences in SI SNP effects between MDD cases and controls.

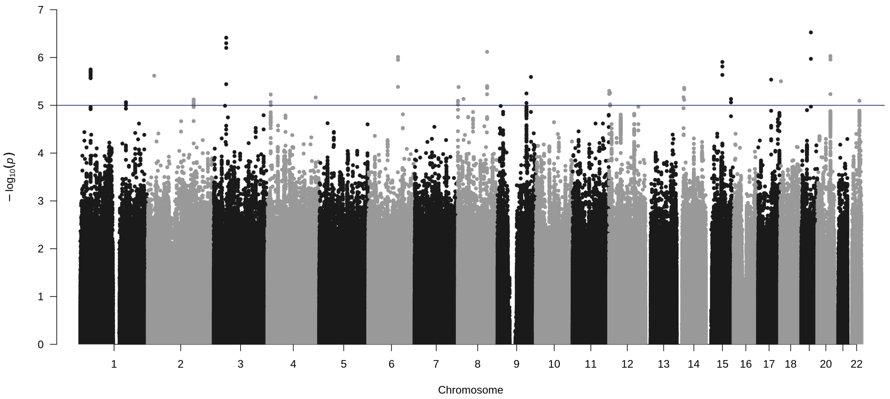

Figure S55: Testing differences in SI SNP effects between PSYCH cases and controls. Blue line indicates $p$-diff < 1x10$^{-5}$. Red line indicates $p$-diff < 5x10$^{-8}$.

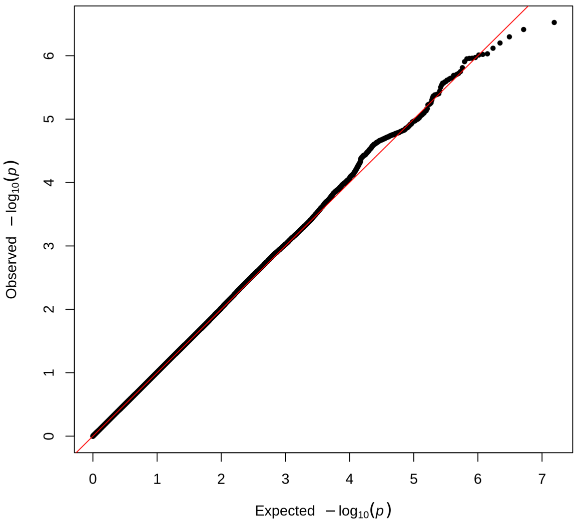

Figure S56: QQ plot showing the distribution of $p$-diff when testing for differences in SI SNP effects between PSYCH cases and controls.

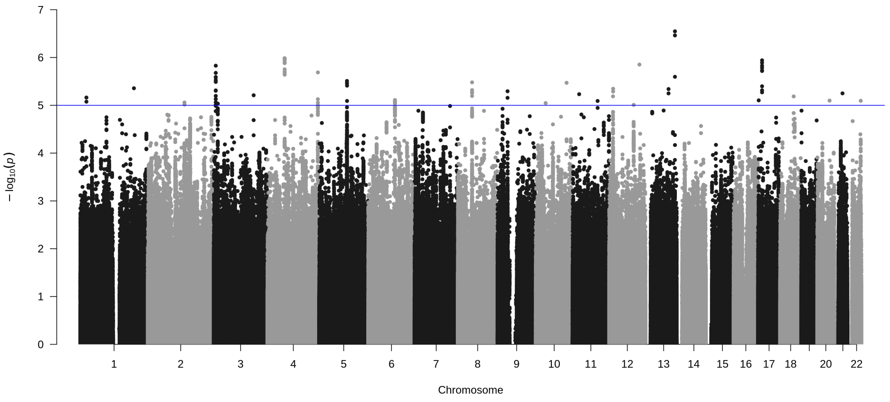

Figure S57: Testing differences in SI SNP effects between sexes. Blue line indicates $p$-diff $< 1x10^{-5}$. Red line indicates $p$-diff $< 5x10^{-8}$.

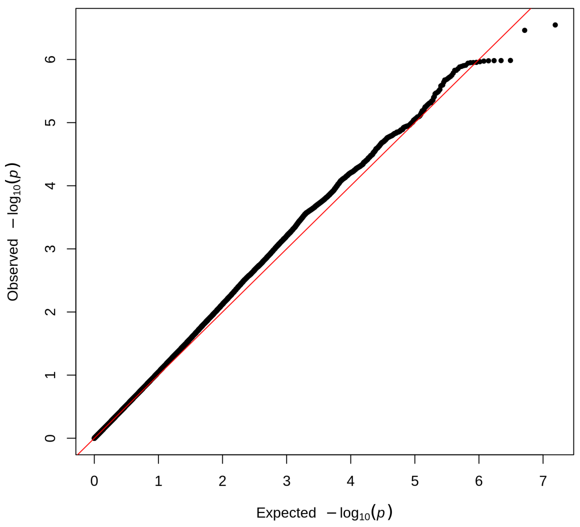

Figure S58: QQ plot showing the distribution of $p$-diff when testing for differences in SI SNP effects between sexes.

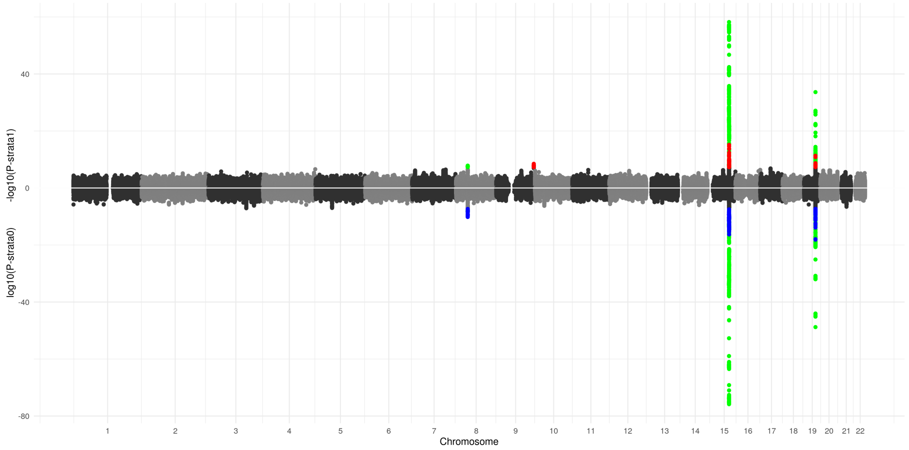

Figure S59: Miami plot for CPD (binned) between late-onset and early-onset smokers. *P*-values for late-onset smokers are shown above the x-axis and *p*-values for early-onset smokers are shown below the x-axis. Red and blue SNPs reached *p*-value < 5x10$^{-8}$ in one strata without reaching 1x10$^{-5}$ in the opposing strata. Green SNPs reached *p*-value < 5x10$^{-8}$ in both strata.

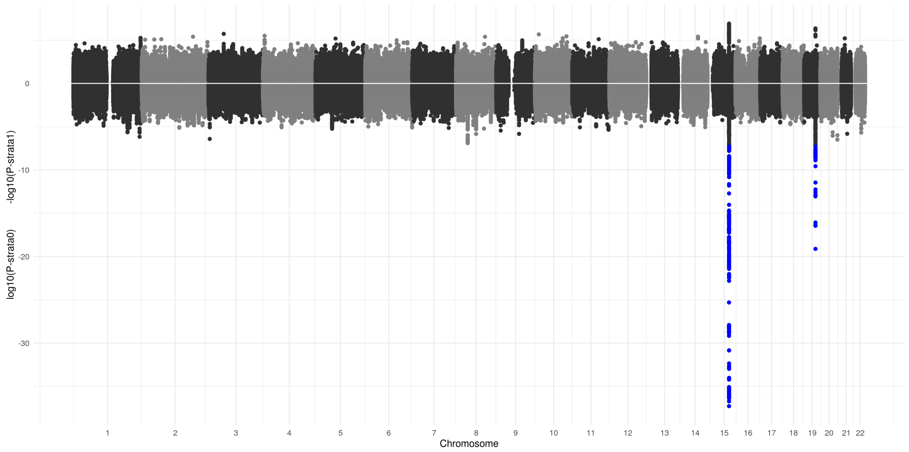

Figure S60: Miami plot for CPD (binned) between GAD cases and controls. *P*-values for GAD cases are shown above the x-axis and *p*-values for GAD controls are shown below the x-axis. Red and blue SNPs reached *p*-value < 5x10$^{-8}$ in one strata without reaching 1x10$^{-5}$ in the opposing strata. Green SNPs reached *p*-value < 5x10$^{-8}$ in both strata.

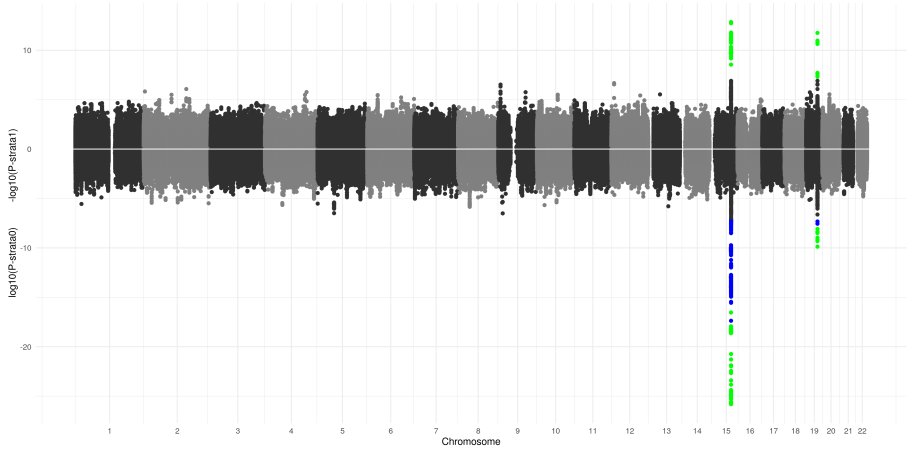

Figure S61: Miami plot for CPD (binned) between MDD cases and controls. $P$-values for MDD cases are shown above the x-axis and $p$-values for MDD controls are shown below the x-axis. Red and blue SNPs reached $p$-value $< 5x10^{-8}$ in one strata without reaching $1x10^{-5}$ in the opposing strata. Green SNPs reached $p$-value $< 5x10^{-8}$ in both strata.

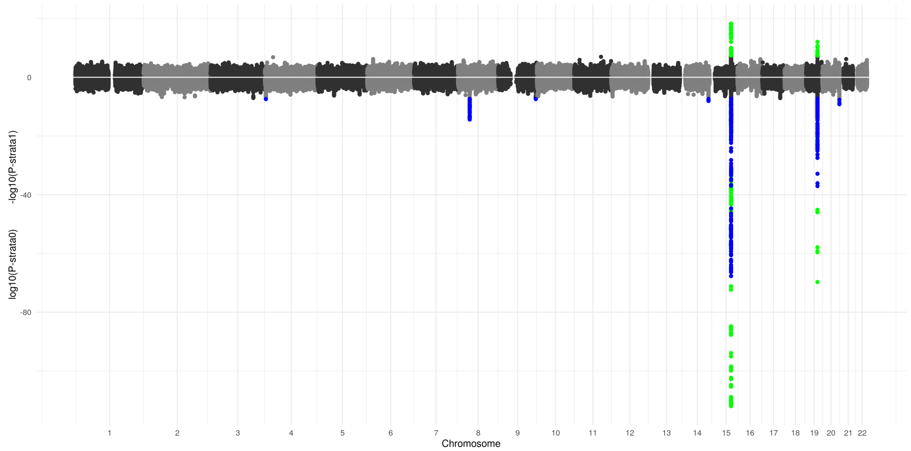

Figure S62: Miami plot for CPD (binned) between PSYCH cases and controls. $P$-values for PSYCH cases are shown above the x-axis and $p$-values for PSYCH controls are shown below the x-axis. Red and blue SNPs reached $p$-value $<$ $5x10^{-8}$ in one strata without reaching $1x10^{-5}$ in the opposing strata. Green SNPs reached $p$-value $< 5x10^{-8}$ in both strata.

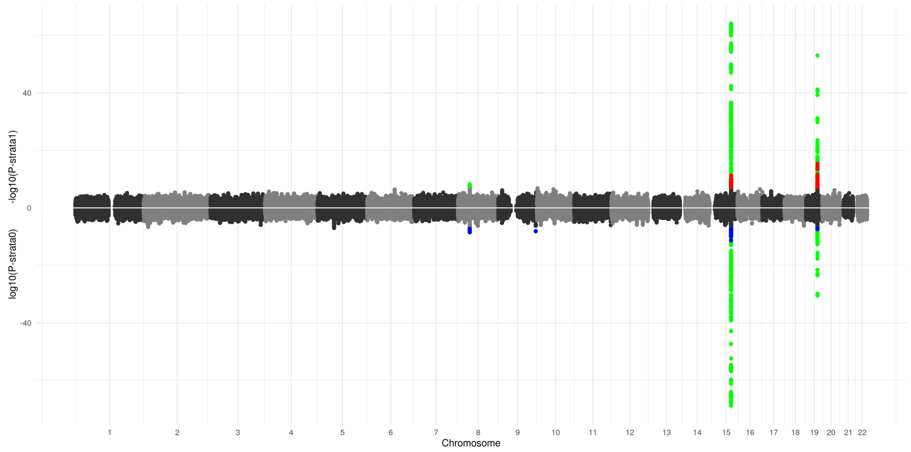

Figure S63: Miami plot for CPD (binned) between sexes. $P$-values for males are shown above the x-axis and $p$-values for females are shown below the x-axis. Red and blue SNPs reached $p$-value $< 5x10^{-8}$ in one strata without reaching $1x10^{-5}$ in the opposing strata. Green SNPs reached $p$-value $< 5x10^{-8}$ in both strata.

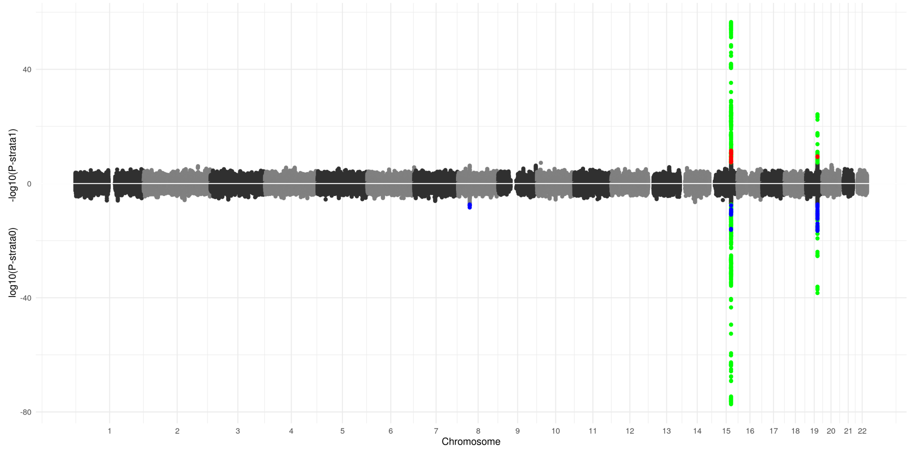

Figure S64: Miami plot for CPD (dichotomized) between late-onset and early-onset smokers. $P$-values for late-onset smokers are shown above the x-axis and $p$-values for early-onset smokers are shown below the x-axis. Red and blue SNPs reached $p$-value $< 5x10^{-8}$ in one strata without reaching $1x10^{-5}$ in the opposing strata. Green SNPs reached $p$-value $< 5x10^{-8}$ in both strata.

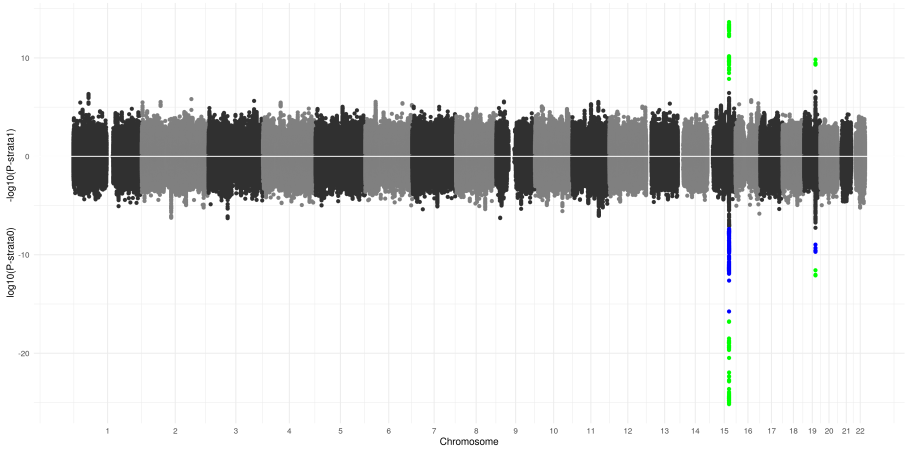

Figure S65: Miami plot for CPD (dichotomized) between MDD cases and controls. $P$-values for MDD cases are shown above the x-axis and $p$-values for MDD controls are shown below the x-axis. Red and blue SNPs reached $p$-value $< 5x10^{-8}$ in one strata without reaching $1x10^{-5}$ in the opposing strata. Green SNPs reached $p$-value $< 5x10^{-8}$ in both strata.

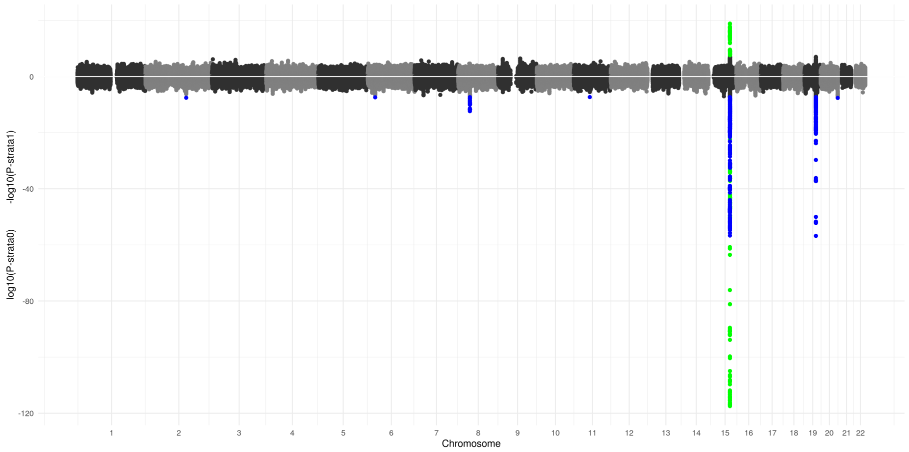

Figure S66: Miami plot for CPD (dichotomized) between PSYCH cases and controls. $P$-values for PSYCH cases are shown above the x-axis and $p$-values for PSYCH controls are shown below the x-axis. Red and blue SNPs reached $p$-value $< 5x10^{-8}$ in one strata without reaching $1x10^{-5}$ in the opposing strata. Green SNPs reached $p$-value $< 5x10^{-8}$ in both strata.

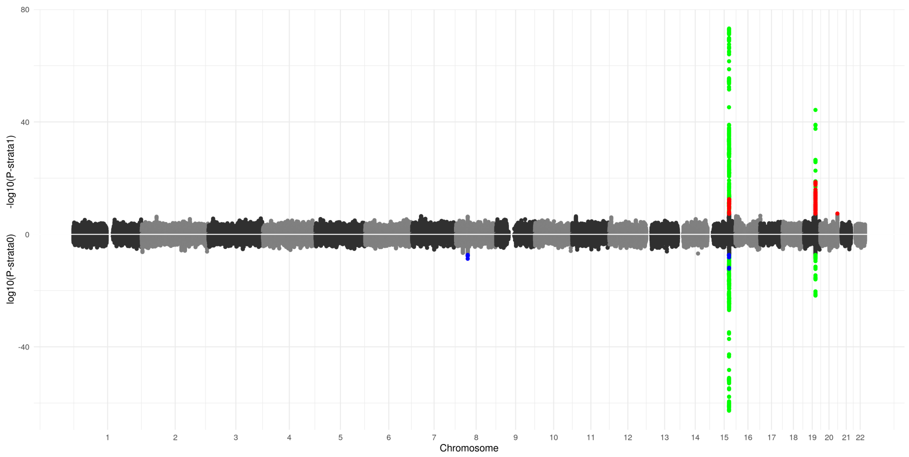

Figure S67: Miami plot for CPD (dichotomized) between sexes. $P$-values for males are shown above the x-axis and $p$-values for females are shown below the x-axis. Red and blue SNPs reached $p$-value $< 5\text{x}10^{-8}$ in one strata without reaching $1\text{x}10^{-5}$ in the opposing strata. Green SNPs reached $p$-value $< 5\text{x}10^{-8}$ in both strata.

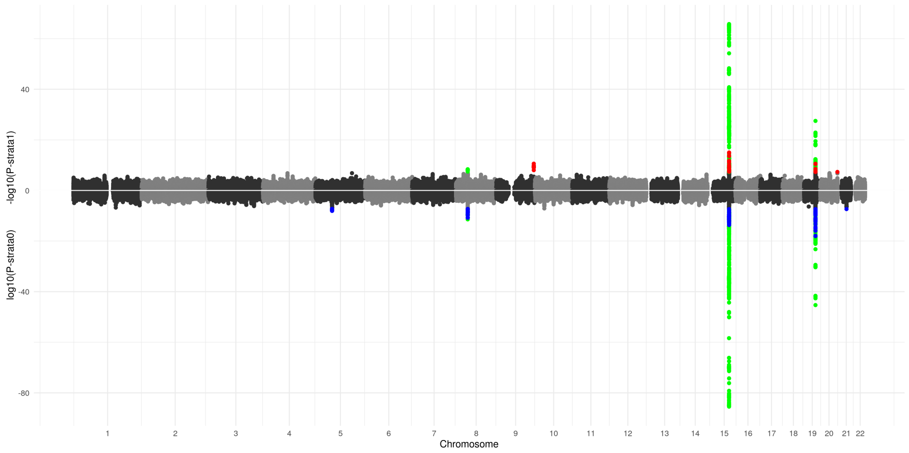

Figure S68: Miami plot for CPD (log transformed) between late-onset and early-onset smokers. $P$-values for late-onset smokers are shown above the x-axis and $p$-values for early-onset smokers are shown below the x-axis. Red and blue SNPs reached $p$-value $< 5\text{x}10^{-8}$ in one strata without reaching $1\text{x}10^{-5}$ in the opposing strata. Green SNPs reached $p$-value $< 5\text{x}10^{-8}$ in both strata.

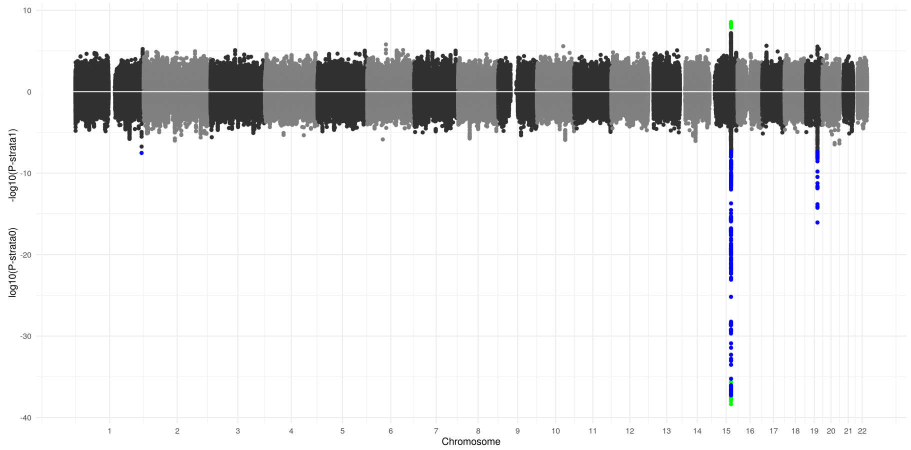

Figure S69: Miami plot for CPD (log transformed) between GAD cases and controls. *P*-values for GAD cases are shown above the x-axis and *p*-values for GAD controls are shown below the x-axis. Red and blue SNPs reached *p*-value $< 5x10^{-8}$ in one strata without reaching $1x10^{-5}$ in the opposing strata. Green SNPs reached *p*-value $< 5x10^{-8}$ in both strata.

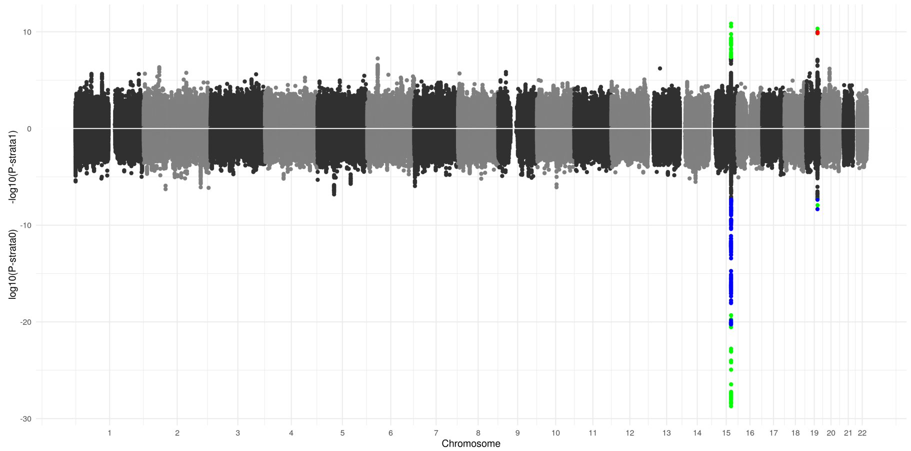

Figure S70: Miami plot for CPD (log transformed) between MDD cases and controls. *P*-values for MDD cases are shown above the x-axis and *p*-values for MDD controls are shown below the x-axis. Red and blue SNPs reached *p*-value $< 5x10^{-8}$ in one strata without reaching $1x10^{-5}$ in the opposing strata. Green SNPs reached *p*-value $< 5x10^{-8}$ in both strata.

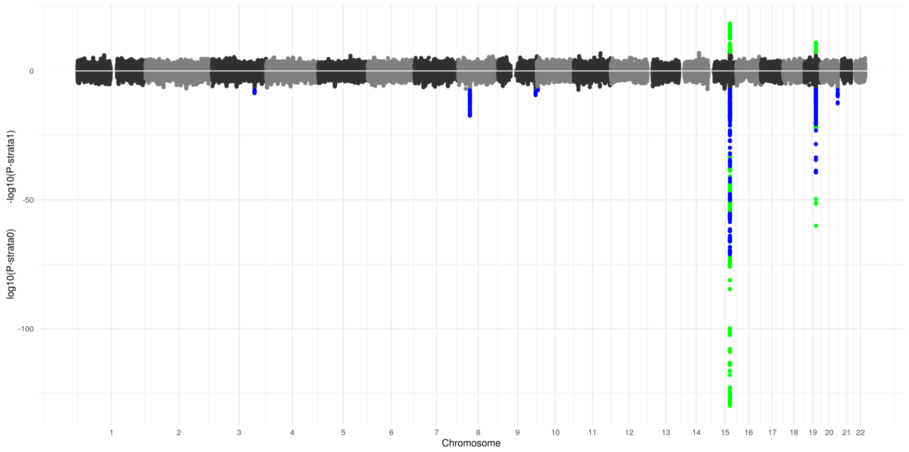

Figure S71: Miami plot for CPD (log transformed) between PSYCH cases and controls. $P$-values for PSYCH cases are shown above the x-axis and $p$-values for PSYCH controls are shown below the x-axis. Red and blue SNPs reached $p$-value $< 5x10^{-8}$ in one strata without reaching $1x10^{-5}$ in the opposing strata. Green SNPs reached $p$-value $< 5x10^{-8}$ in both strata.

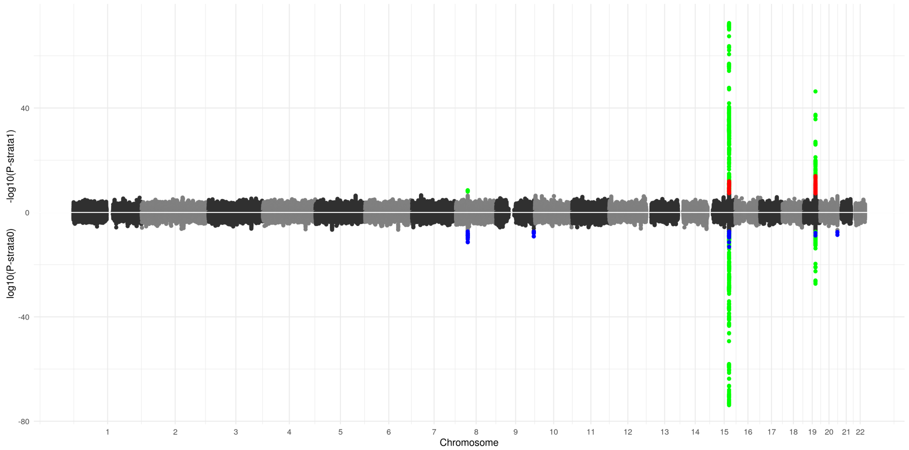

Figure S72: Miami plot for CPD (log transformed) between sexes. $P$-values for males are shown above the x-axis and $p$-values for females are shown below the x-axis. Red and blue SNPs reached $p$-value $< 5x10^{-8}$ in one strata without reaching $1x10^{-5}$ in the opposing strata. Green SNPs reached $p$-value $< 5x10^{-8}$ in both strata.

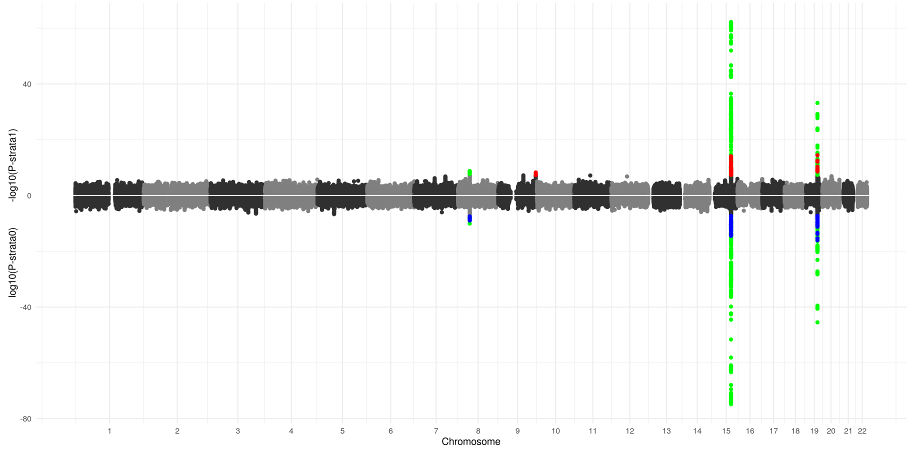

Figure S73: Miami plot for CPD (raw scale) between late-onset and early-onset smokers. *P*-values for late-onset smokers are shown above the x-axis and *p*-values for early-onset smokers are shown below the x-axis. Red and blue SNPs reached *p*-value < 5x10$^{-8}$ in one strata without reaching 1x10$^{-5}$ in the opposing strata. Green SNPs reached *p*-value < 5x10$^{-8}$ in both strata.

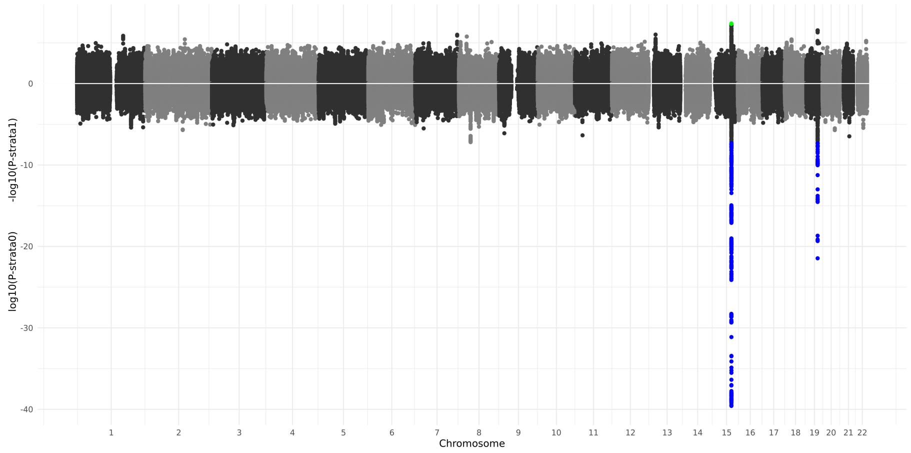

Figure S74: Miami plot for CPD (raw scale) between GAD cases and controls. *P*-values for GAD cases are shown above the x-axis and *p*-values for GAD controls are shown below the x-axis. Red and blue SNPs reached *p*-value < 5x10$^{-8}$ in one strata without reaching 1x10$^{-5}$ in the opposing strata. Green SNPs reached *p*-value < 5x10$^{-8}$ in both strata.

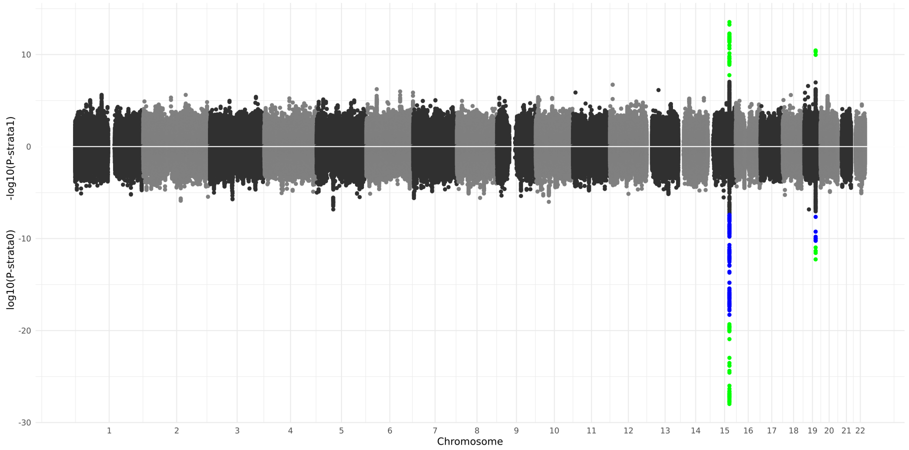

Figure S75: Miami plot for CPD (raw scale) between MDD cases and controls. $P$-values for MDD cases are shown above the x-axis and $p$-values for MDD controls are shown below the x-axis. Red and blue SNPs reached $p$-value $<$ $5 \times 10^{-8}$ in one strata without reaching $1 \times 10^{-5}$ in the opposing strata. Green SNPs reached $p$-value $< 5 \times 10^{-8}$ in both strata.

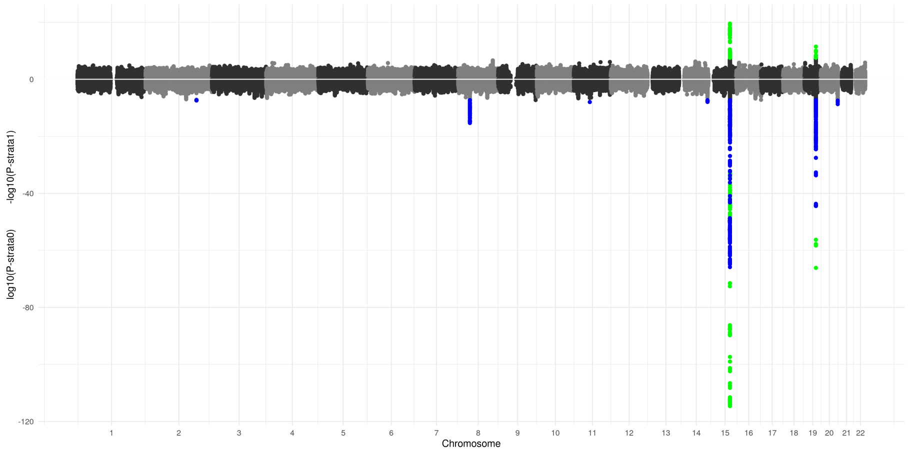

Figure S76: Miami plot for CPD (raw scale) between PSYCH cases and controls. $P$-values for PSYCH cases are shown above the x-axis and $p$-values for PSYCH controls are shown below the x-axis. Red and blue SNPs reached $p$-value $<$ $5 \times 10^{-8}$ in one strata without reaching $1 \times 10^{-5}$ in the opposing strata. Green SNPs reached $p$-value $< 5 \times 10^{-8}$ in both strata.

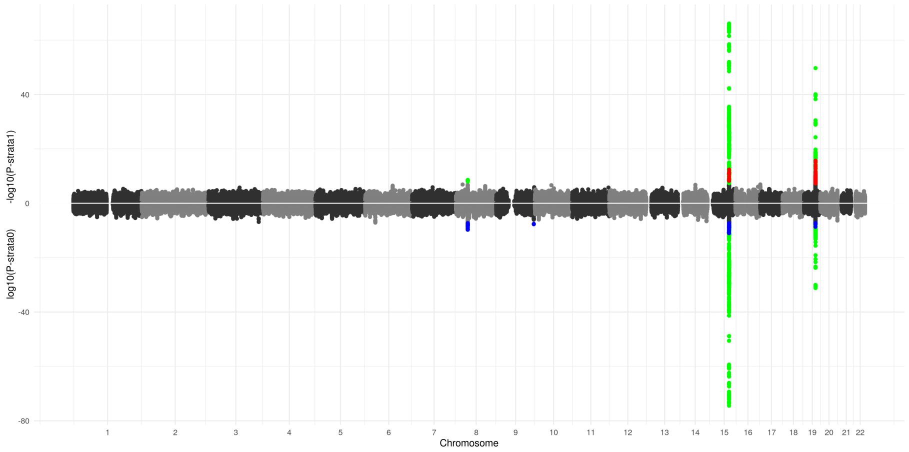

Figure S77: Miami plot for CPD (raw scale) between sexes. $P$-values for males are shown above the x-axis and $p$-values for females are shown below the x-axis. Red and blue SNPs reached $p$-value $<$ 5x10$^{-8}$ in one strata without reaching 1x10$^{-5}$ in the opposing strata. Green SNPs reached $p$-value $<$ 5x10$^{-8}$ in both strata.

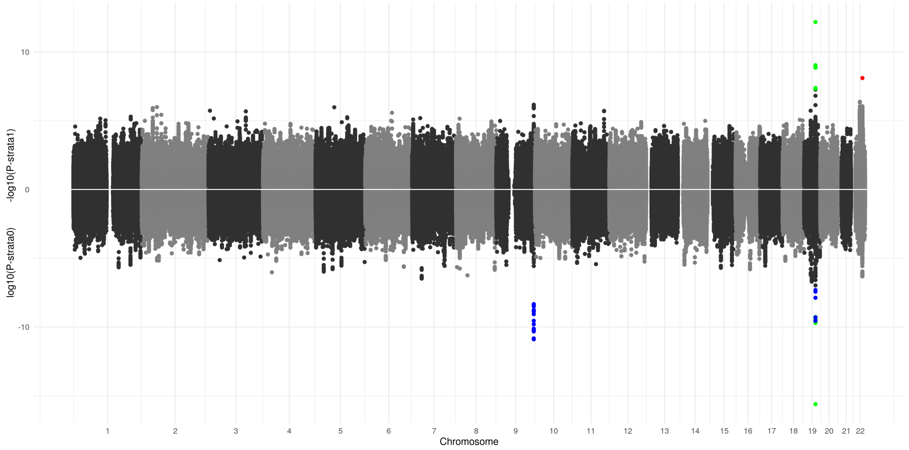

Figure S78: Miami plot for SC between late-onset and early-onset smokers. $P$-values for late-onset smokers are shown above the x-axis and $p$-values for early-onset smokers are shown below the x-axis. Red and blue SNPs reached $p$-value $<$ 5x10$^{-8}$ in one strata without reaching 1x10$^{-5}$ in the opposing strata. Green SNPs reached $p$-value $<$ 5x10$^{-8}$ in both strata.

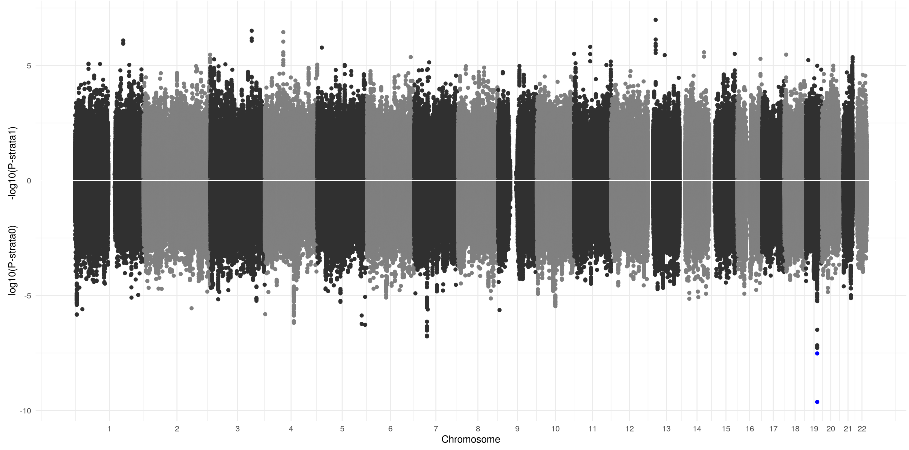

Figure S79: Miami plot for SC between GAD cases and controls. $P$-values for GAD cases are shown above the x-axis and $p$-values for GAD controls are shown below the x-axis. Red and blue SNPs reached $p$-value $< 5x10^{-8}$ in one strata without reaching $1x10^{-5}$ in the opposing strata. Green SNPs reached $p$-value $<$ $5x10^{-8}$ in both strata.

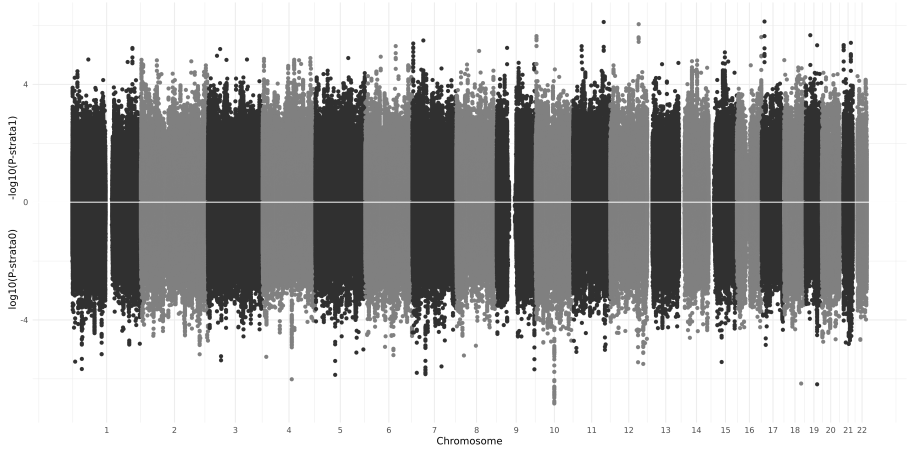

Figure S80: Miami plot for SC between MDD cases and controls. $P$-values for MDD cases are shown above the x-axis and $p$-values for MDD controls are shown below the x-axis. Red and blue SNPs reached $p$-value $< 5x10^{-8}$ in one strata without reaching $1x10^{-5}$ in the opposing strata. Green SNPs reached $p$-value $<$ $5x10^{-8}$ in both strata.

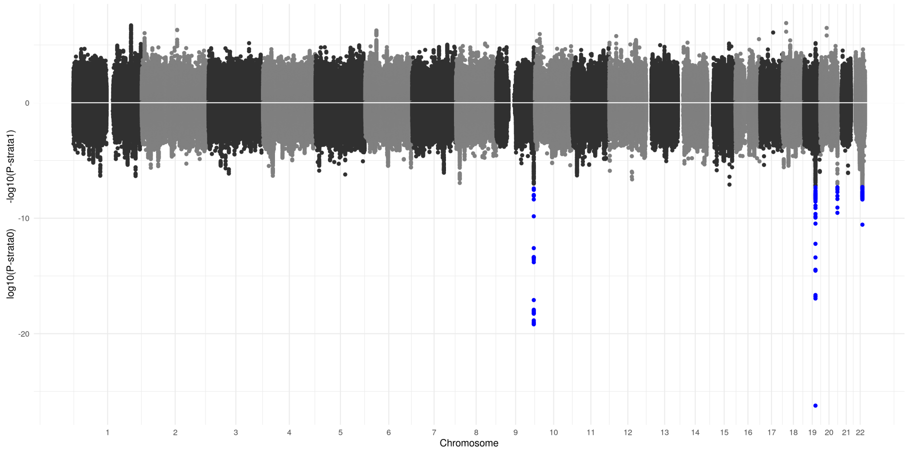

Figure S81: Miami plot for SC between PSYCH cases and controls. *P*-values for PSYCH cases are shown above the x-axis and *p*-values for PSYCH controls are shown below the x-axis. Red and blue SNPs reached *p*-value < 5x10$^{-8}$ in one strata without reaching 1x10$^{-5}$ in the opposing strata. Green SNPs reached *p*-value < 5x10$^{-8}$ in both strata.

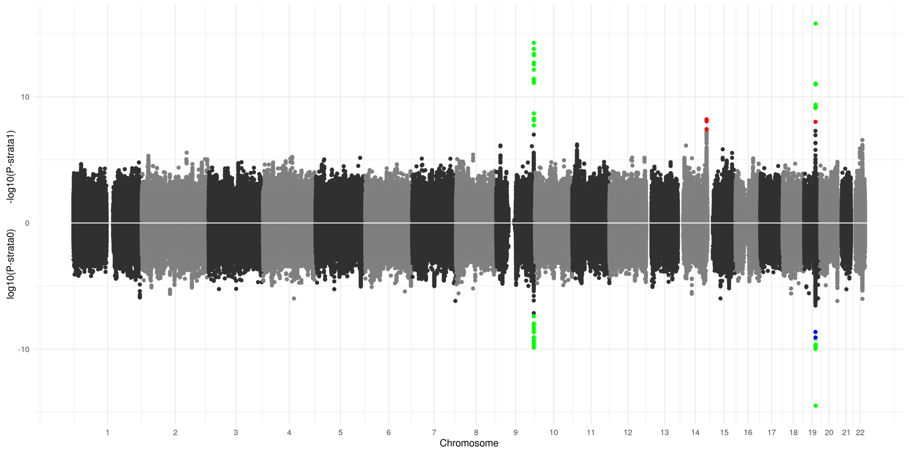

Figure S82: Miami plot for SC between sexes. *P*-values for males are shown above the x-axis and *p*-values for females are shown below the x-axis. Red and blue SNPs reached *p*-value < 5x10$^{-8}$ in one strata without reaching 1x10$^{-5}$ in the opposing strata. Green SNPs reached *p*-value < 5x10$^{-8}$ in both strata.

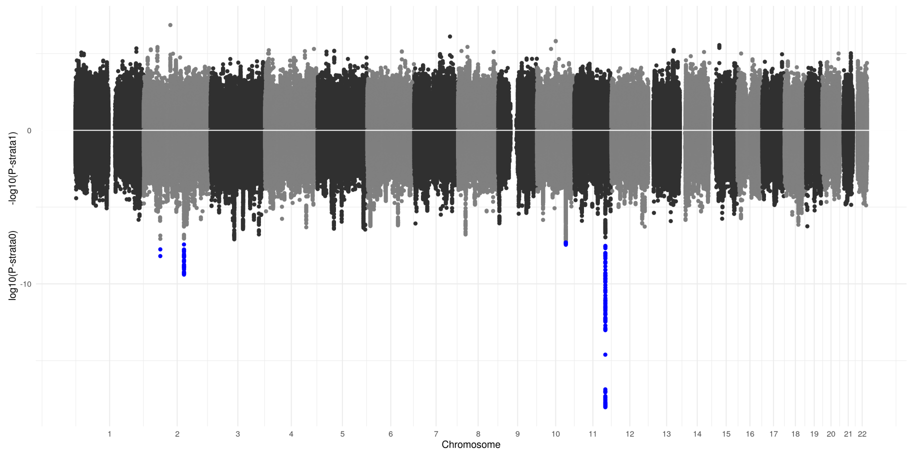

Figure S83: Miami plot for SI between GAD cases and controls. $P$-values for GAD cases are shown above the x-axis and $p$-values for GAD controls are shown below the x-axis. Red and blue SNPs reached $p$-value $< 5x10^{-8}$ in one strata without reaching $1x10^{-5}$ in the opposing strata. Green SNPs reached $p$-value $<$ $5x10^{-8}$ in both strata.

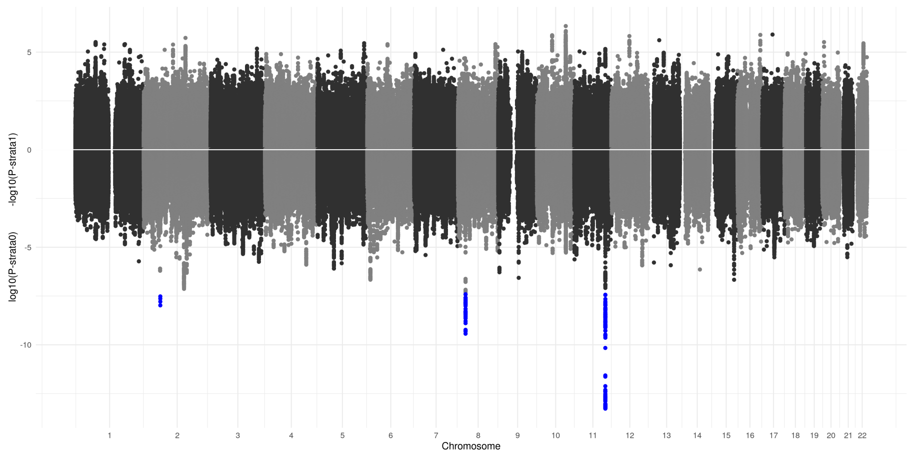

Figure S84: Miami plot for SI between MDD cases and controls. $P$-values for MDD cases are shown above the x-axis and $p$-values for MDD controls are shown below the x-axis. Red and blue SNPs reached $p$-value $< 5x10^{-8}$ in one strata without reaching $1x10^{-5}$ in the opposing strata. Green SNPs reached $p$-value $<$ $5x10^{-8}$ in both strata.

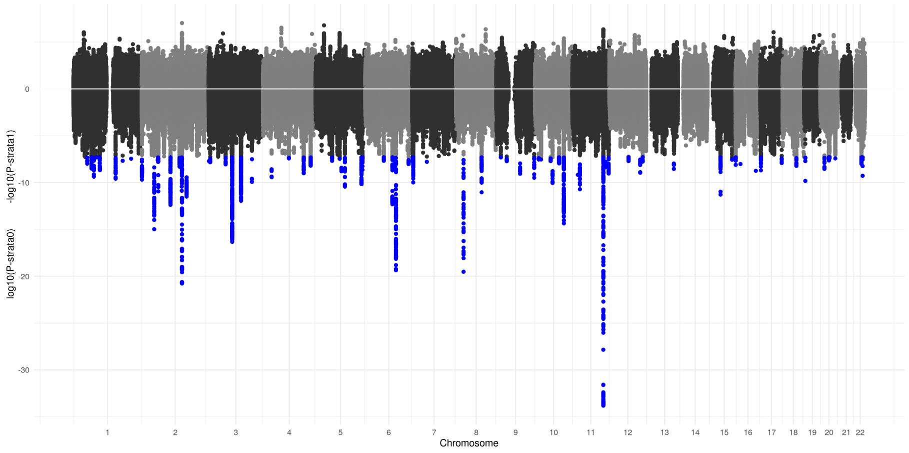

Figure S85: Miami plot for SI between PSYCH cases and controls. $P$-values for PSYCH cases are shown above the x-axis and $p$-values for PSYCH controls are shown below the x-axis. Red and blue SNPs reached $p$-value $< 5x10^{-8}$ in one strata without reaching $1x10^{-5}$ in the opposing strata. Green SNPs reached $p$-value $< 5x10^{-8}$ in both strata.

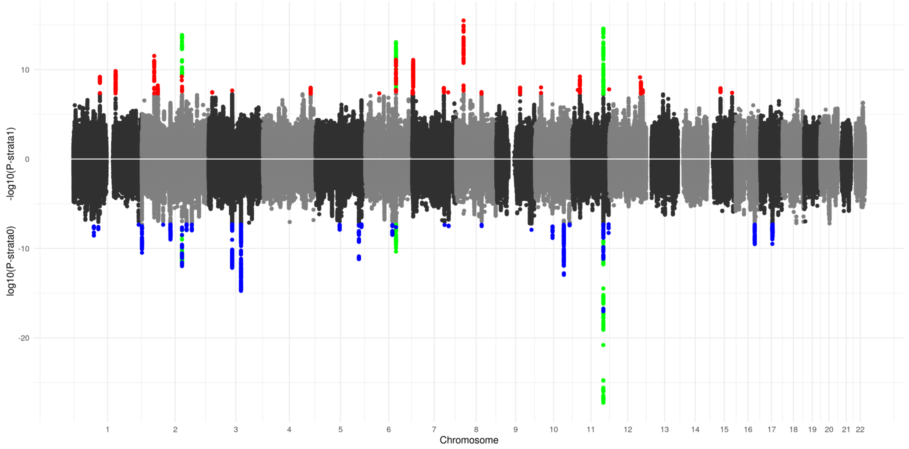

Figure S86: Miami plot for SI between sexes. $P$-values for males are shown above the x-axis and $p$-values for females are shown below the x-axis. Red and blue SNPs reached $p$-value $< 5x10^{-8}$ in one strata without reaching $1x10^{-5}$ in the opposing strata. Green SNPs reached $p$-value $< 5x10^{-8}$ in both strata.

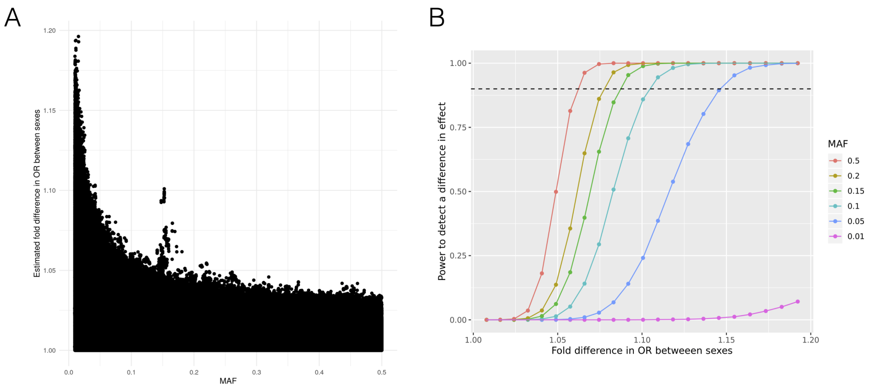

Figure S87: Observed sex differences in SNP effects with simulations to detect differences. (A) Comparing estimated differences in sex-specific SNP effects against minor allele frequency. Shown is the MAF obtained in females, with the correlation between male and female MAF $> 0.99$, genome-wide. The estimated fold difference in OR between sexes was obtained by taking the maximum between $\hat{OR}_{male}/\hat{OR}_{female}$ and $\hat{OR}_{female}/\hat{OR}_{male}$ in order to obtain a fold difference that is always greater than one. The peak at MAF $= 0.15$ is localized to the MHC region. (B) Power to detect a difference in effects at genome-wide significance. Power was estimated from 5000 simulation replicates. In all simulations, the female-specific effect was kept constant, while varying the male-specific effect. The dashed line is at 90% power.

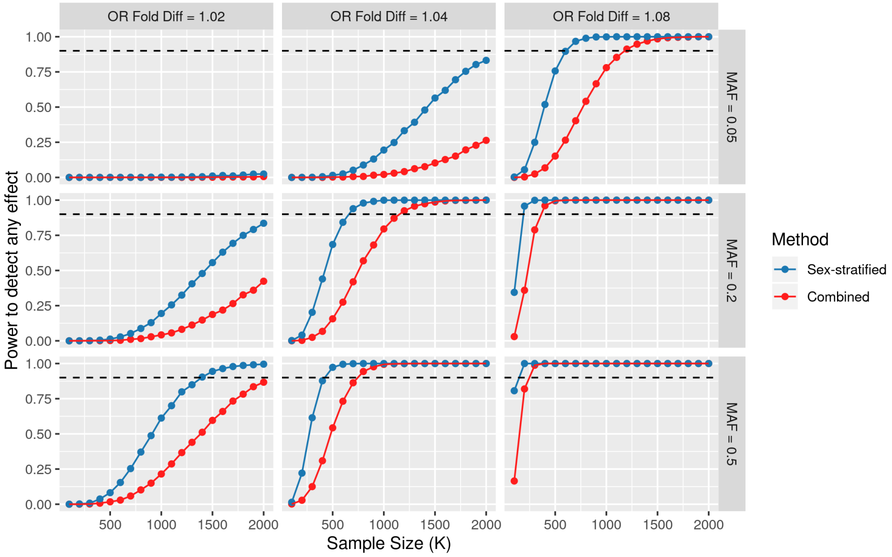

Figure S88: Power to detect any effect (whether it affects males, females, or both) at genome-wide significance (5x10$^{-8}$), across degrees of SNP effect heterogeneity and MAF. Total sample size (all males and females; x-axis) varied, while keeping the male fraction (0.45), male SI case fraction (0.51), and female SI case fraction (0.39) constant. In blue, sex-stratified regression was used, and the minimum between p-female and p-male is used to estimate power. In red, sex-combined regression was used, while including sex as a covariate. The degree of SNP effect heterogeneity (vertical facets) is expressed as the male-specific OR divided by the female-specific OR. In all simulations, the female-specific effect was kept constant, while varying the male-specific effect. Power was estimated using 5000 simulation replicates. The dashed line marks 90% power.

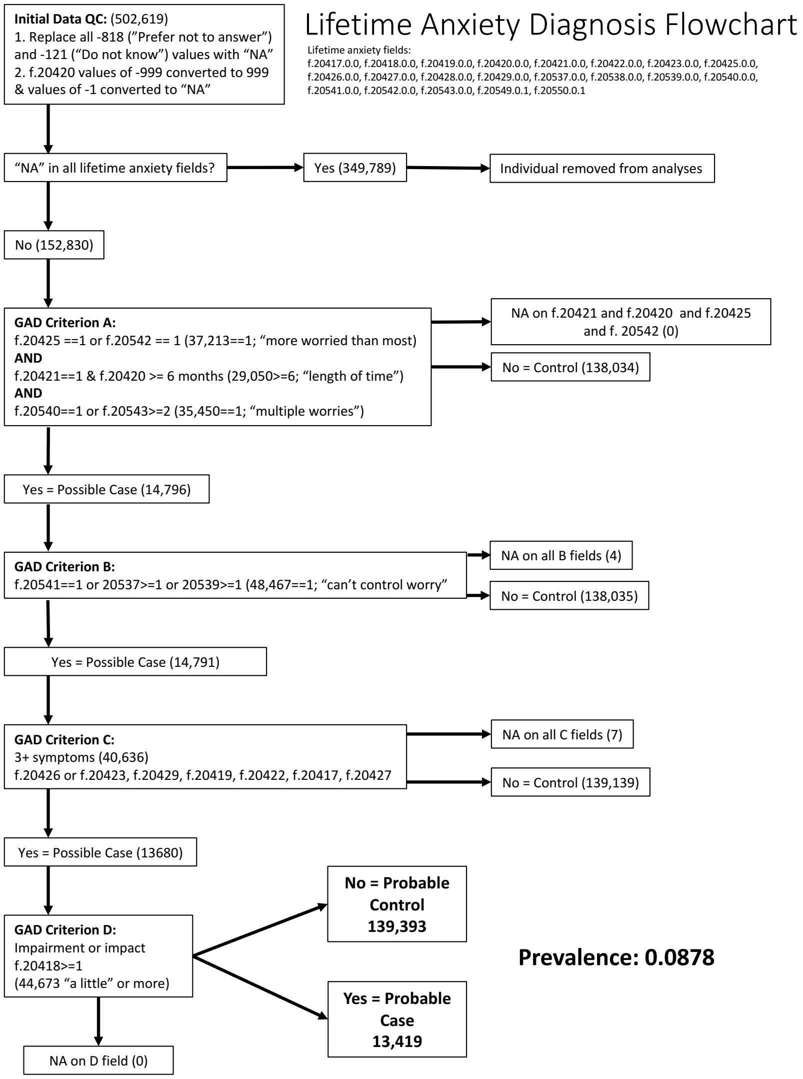

Figure S89: Flowchart for GAD DSM-V-like assignment
